# Aversive memory formation in humans is determined by an amygdala-hippocampus phase code

**DOI:** 10.1101/2021.09.06.21262859

**Authors:** Manuela Costa, Diego Lozano-Soldevilla, Antonio Gil-Nagel, Rafael Toledano, Carina Oehrn, Lukas Kunz, Mar Yebra, Costantino Mendez-Bertolo, Lennart Stieglitz, Johannes Sarnthein, Nikolai Axmacher, Stephan Moratti, Bryan A. Strange

## Abstract

Memory for aversive events is central to survival, but can also become maladaptive in psychiatric disorders. Emotional memory relies on the amygdala and hippocampus, but the neural dynamics of their communication during emotional memory encoding remain unknown. Using simultaneous intracranial recordings from both structures in human patients, we show that in response to emotionally aversive, but not neutral, visual stimuli, the amygdala transmits unidirectional influence on the hippocampus through theta oscillations. Critically, successful emotional memory encoding depends on the precise amygdala theta phase to which hippocampal gamma activity and neuronal firing couple. The phase difference between subsequently remembered *vs*. not-remembered emotional stimuli translates to ∼25-45 milliseconds, a time period that enables lagged coherence between amygdala and downstream hippocampal gamma activity. These results reveal a mechanism whereby amygdala theta phase coordinates transient coherence between amygdala and hippocampal gamma activity to facilitate the encoding of aversive memories in humans.

## The phase of human amygdala theta to which hippocampal gamma activity and neuronal firing couples determines subsequent remembering of aversive events

We tend to remember emotional events better than neutral ones. Although adaptive to survival, emotional memory enhancement for traumatic experience can contribute to anxiety (*1*) and post-traumatic stress disorders (*2*). Research in animal models and humans implicate the amygdala (*3, 4*), and hippocampus (*5, 6*) in emotional memory. Patients with selective amygdala lesions show reduced episodic memory for emotional items (*7–9*) and functional MRI (fMRI) studies have reported increased amygdala responses to emotional, relative to neutral, stimuli during memory encoding (*10–12*). However, animal (*13*) and humans studies (*10–12, 14*) suggest the amygdala is not a site of long-term episodic memory storage, but rather that it influences memory storage processes in the hippocampus. Despite this long-standing modulation hypothesis (*13*), the circuit-level neurophysiological mechanism underlying amygdalo-hippocampal interactions is not yet understood. One proposal is that the interplay between the two structures occurs via coordinated oscillatory activity (*15, 16*). In humans, limitations of current non-invasive neuroimaging techniques dictate that this mechanism be determined using direct electrophysiological recordings from both amygdala and hippocampus. We therefore recorded intracranially from two cohorts of pharmaco-resistant epilepsy patients (cohort 1: *n=*13, Table S1; cohort 2: *n*=6, Table S2) while they performed an emotional memory task (**Fig. 1A**). In cohort 1, electrodes had been implanted in the amygdala and in 8 patients also in the ipsilateral hippocampus (**Fig. 1C**), allowing simultaneous recordings of oscillatory responses associated with emotional stimulus encoding from both structures (fig S1A), and, critically, the assessment of emotional memory-dependent amygdala-hippocampal connectivity. Patients viewed neutral and emotional complex scenes and 24 h later performed a recognition memory test, during which these scenes were presented again, intermixed with novel emotional and neutral foils. Patients made ‘remember’, ‘know’, and ‘new’ (R, K, N) responses to distinguish memories accompanied by a sense of recollection (R) rather than familiarity (K) (*17*). Emotion selectively enhances memories accompanied by a sense of recollection (*18*), and previous fMRI data show amygdala encoding-related responses are larger to subsequently recollected *vs*. familiar or forgotten emotional stimuli (*11*).

**Fig. 1.**
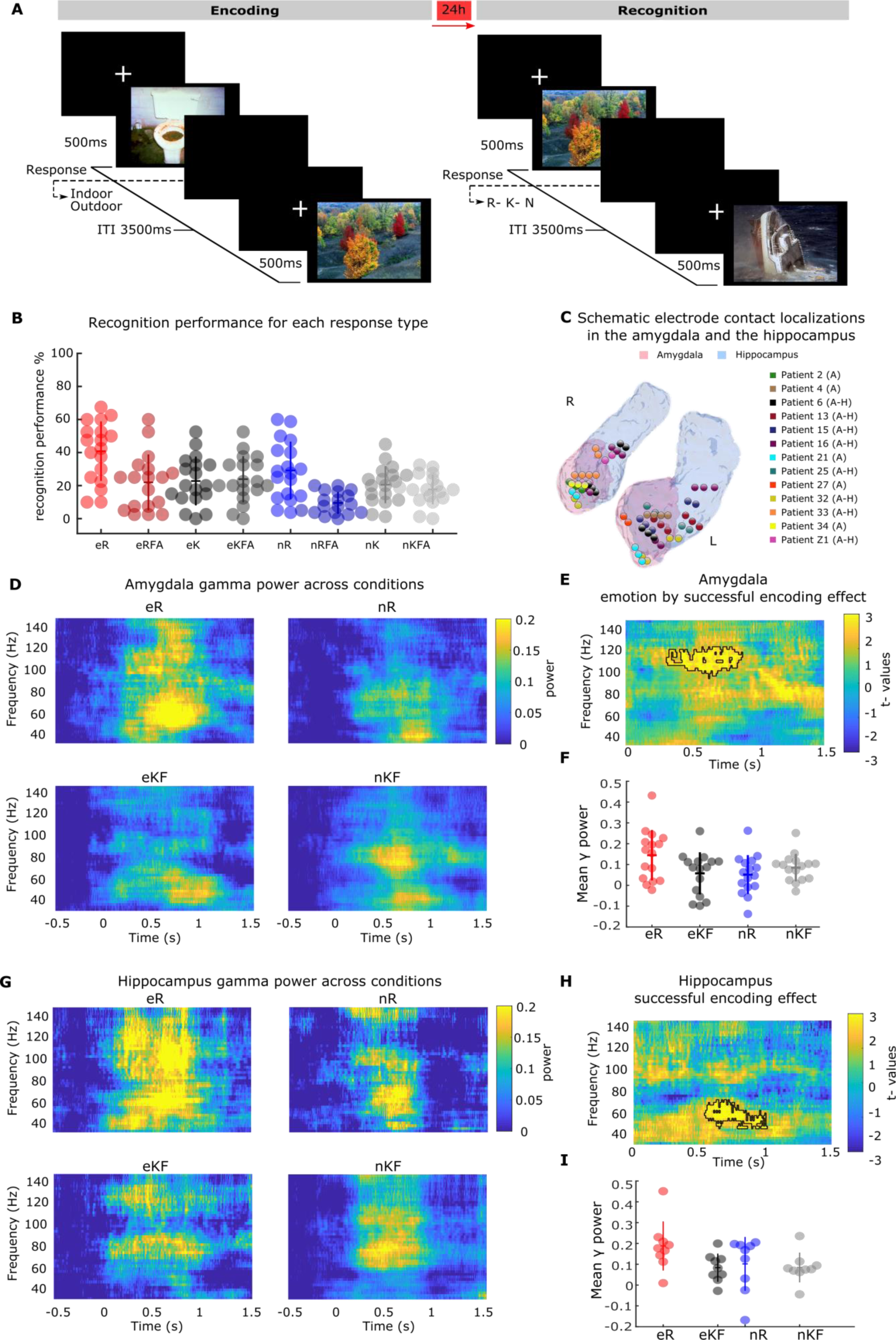
Fast amygdala gamma activity tracks enhanced emotional memory formation, whereas hippocampal gamma activity increased during emotional and neutral memory formation. (A) Task design. Examples of aversive and neutral scene presented during encoding and recognition sessions. (B) Recognition performance for emotionally aversive (e) and neutral (n) correctly remembered, remembered false alarms, correctly known and known false alarms (R, RFA, K, KFA, respectively) responses in the full patient cohort (*n=*18). Individual patient performance is plotted, with horizontal/vertical lines indicating the mean/±s.e.m. (C) Summary of electrode contact localization in the left and right amygdala (pink) and hippocampus (light blue) for all patients included in the analysis (*n=*13 patients with a total of 17 electrodes in the amygdala (A), 4 patients have bilateral amygdala electrodes, *n*=8 patients with a total of 9 electrodes in both structures (A-H), 1 patient has bilateral hippocampal electrodes). Amygdala and hippocampus are an average of these structures segmented for each patient. (D) Time-frequency plots of amygdala power change in the gamma range for aversive and neutral subsequently remembered, and not remembered (known/forgotten) trials. Colorbar: power change relative to baseline. (E) Time frequency resolved test statistics for the relative power change comparison between emotion and successful encoding effect. Amygdala power in the gamma range is greater for eR *vs*. eKF relative to nR *vs*. nKF scenes. Colorbar: *t*-values, and the black outline indicates the significant cluster. (F) Mean amygdala gamma power in the significant cluster is plotted for each amygdala electrode. (G) Time-frequency plots of hippocampus power change in the gamma range for aversive and neutral subsequently remembered KF trials. Colorbar: power change relative to baseline. (H) Time frequency resolved test statistics showing the main effect of successful encoding (R vs. KF). Hippocampus power change in the gamma range is greater for subsequently remembered (R) than subsequently known and forgotten trials (KF), regardless of emotion. Colorbar: *t*-values. (I) Mean hippocampus gamma power in the significant cluster.

Across both patient cohorts, recollection performance for emotional stimuli (eR) was higher than all other response categories (**Fig. 1B**). As is commonly observed, R false alarm rate was higher for emotional (mean ± s.e.m.: 21.94% ± 3.98) compared to neutral stimuli (9.51±1.56; t_17_=3.76, *P*=0.002, d=0.878). Performance for familiar responses was equivalent for emotional (22.77±3.48) and neutral items (20.55±2.65; t_17_=1.01, *P*=0.325, d=0.170). Thus, memory recollection performance was evaluated by comparing the rate of R with K responses for emotional (eR, eK) *vs*. neutral items (nR, nK), as described previously (*11*). This confirmed a significant emotion by memory interaction (repeated measures ANOVA *F*_(2, 17)_ =5.47, *P* =0.032, η²=0.244) (**Fig. 1B**; Table S3-S4, fig. S2).

To determine the oscillatory responses associated with enhanced emotional memory encoding, we employed a subsequent memory approach, categorizing trials according to whether patients later remembered (R) the stimulus presented at encoding or not (known/forgotten items, KF) to operationalize successful encoding. In the amygdala (**Fig. 1, D-F**), successful encoding of aversive, but not of neutral scenes, was associated with fast gamma activity (97-125 Hz) starting 0.31 s after stimulus onset (emotion by subsequent memory interaction, summed *t*-value of difference comparison=1012.36, *P*=0.01, **Fig. 1E**; interactions are assessed via *t*-tests between condition differences). Subsequently remembered aversive scenes induced higher gamma power changes than subsequently known/forgotten aversive items (eR *vs.* eKF t_15_=2.12, *P=*0.050, d=0.53, *post-hoc t*-tests on mean power changes across significant time-frequency clusters, **Fig. 1F**). Memory-related responses to neutral stimuli did not show a significant difference (nR *vs.* nKF, t_15_=-1.90, *P*=0.075, d=- 0.47).

Similar amygdala fast gamma responses (time window 0.41–1.1 s) were observed if the Cohort 1 patient group was restricted to those with electrodes in both amygdala and hippocampus (fig. S3). By contrast to the amygdala spectral power changes, hippocampal-induced responses (50-75 Hz from 0.54 s) were associated with subsequent recollection (R *vs*. KF) of both aversive and neutral pictures. (**Fig. 1, G-I**; main effect of successful encoding, summed *t* value=776.82, *P*=0.0035). Regarding lower oscillatory frequencies (1–34 Hz), significant condition differences were not evident in either structure (fig. S4). Note that the power spectral analysis comparing subsequently remembered with either KF responses showed analogous time frequency effects in both amygdala (fig. S5) and hippocampus (fig. S6), with no significant power differences for K *vs*. F in either structure. This supports previous observations showing a selective role of these structures in encoding leading to subsequent recollection as opposed to familiarity responses (*11*), and provides a rationale for collapsing K and F trials in these and subsequent analyses. Note that medial temporal cortical recordings from entorhinal and perirhinal cortex showed greater gamma responses (50-67 Hz) following successful encoding of neutral, but not aversive scenes suggesting a specific mechanism between amygdala and hippocampus for emotional memory formation (fig. S7, *n*=8).

Having characterized amygdala and hippocampal gamma power responses during memory formation, we next systematically examined their coupling during emotional memory formation. In rodents, emotional memory retrieval is associated with amygdalo-hippocampal theta coherence (*15*). In non-human primates, long-range communication between amygdala and cortical regions during aversive learning is achieved via theta coherence, with directional flow of information from amygdala to cortex (*19, 20*). We therefore tested for coherence and directional influence (Granger causality) between the amygdala and hippocampus. Unexpectedly, we did not find emotion-dependent coherence differences across encoding conditions (fig. S8). Directionality analysis using frequency-resolved Granger causality did, however, reveal greater causal influence from amygdala to hippocampus (lag of 0.36 s) during viewing of aversive as compared to neutral scenes. The effect was observed within the theta/alpha band (3-17 Hz) and the time window (0.43 - 0.77 s) in which amygdala gamma activity to subsequently remembered emotional stimuli becomes pronounced (**Fig. 2A-B**; main effect of emotion, summed *t* value=1103.47, *P=*0.0039; main effect of emotion in the opposite direction was not significant (**Fig. 2C**, fig. S9).

**Fig. 2.**
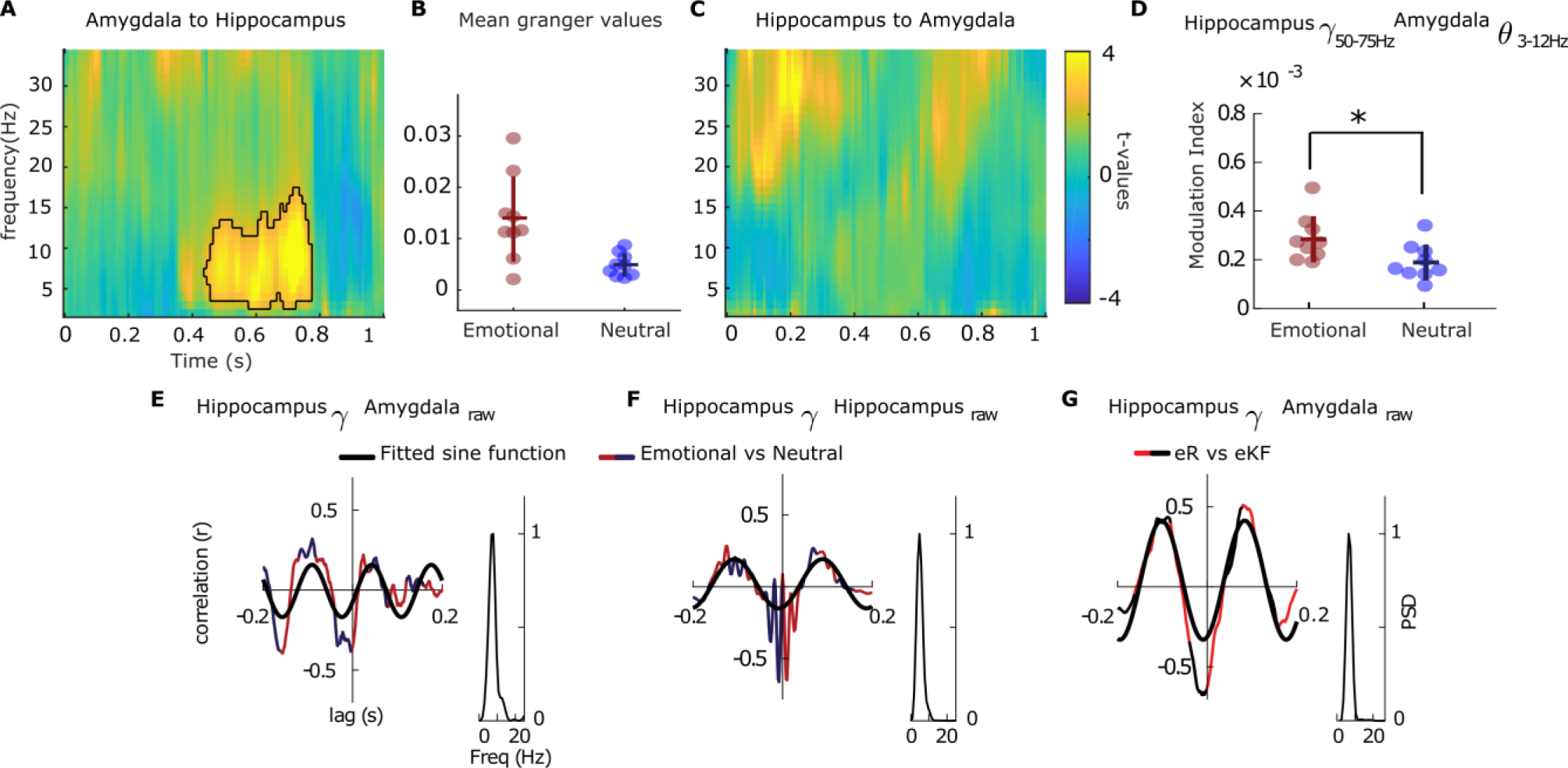
Emotion-dependent amygdala-hippocampus theta-gamma coupling. **(A)** Granger causal influence of amygdala on hippocampal oscillations for aversive *vs*. neutral scenes (main effect of emotion). **(B)** Mean Granger values for the significant cluster (0.43-0.77 s post stimulus onset in the theta/alpha range from 3-17 Hz) is plotted for each amygdala-hippocampus electrode pair (8 patients and a total of 9 electrode pairs). Horizontal/vertical bars represent mean/±s.e.m. **(C)** As for A, but in the direction hippocampus to amygdala. (**D**) Amygdala theta phase (3–12 Hz) to hippocampus gamma amplitude (50–75 Hz) coupling (MI) is greater in response to emotional *vs*. neutral stimuli. Data points represent each pair of electrode contacts located in amygdala and hippocampus. Horizontal/vertical bars represent mean/ ± s.e.m. **(E)** Inter-regional cross-correlation between emotional and neutral (dark blue/dark red) peak trigger average (PTA) (left) and the corresponding power spectral density (PSD) of the cross-correlogram (right) for Patient 16. A theta component is evident in the PSD and fitted sine wave (thick black line, here and in subsequent panels). **(F)** Same representative patient, within-hippocampus cross-correlation between emotional and neutral (blue/red) experimental conditions. PTAs were computed as in **(E)** but averaging the raw local field potentials from the hippocampal recordings. (**G**) Same representative patient, PTA for aversive remember *vs*. know/forgotten (red/black) trials using the peaks of hippocampus gamma activity to average amygdala raw recordings.

The degree to which single neuron spiking phase-locks to human hippocampal theta oscillations is a predictor of memory strength (*21, 22*). Given that spiking has been shown to correlate with gamma power (*23, 24*), we hypothesized that a stronger modulation of hippocampal gamma band activity by amygdala theta oscillations would occur during encoding of aversive scenes leading to remembering. We thus tested the phase-amplitude coupling (PAC) associated with subsequently remembered *vs*. non-remembered emotional items by calculating the modulation index (MI) (*25*) (fig. S10-S13). Amygdala theta phase coupling with hippocampal gamma amplitude was stronger for emotional compared to neutral scenes (F_(1, 8)_=6.73, *P=*0.031, η^2^=0.457, **Fig. 2D**), similar to previous observations during passive viewing of emotional (fearful) faces *vs*. neutral landscapes (*16*). However, PAC was unrelated to memory formation (*i.e.*, MI values did not show a significant emotion by memory interaction).

This result was further corroborated using a complementary approach which isolated hippocampal gamma bursts during successful and unsuccessful encoding of aversive scenes and used them to trigger averages in amygdala raw field potentials. The cross-correlogram (CC) obtained from the peak-triggered averages (PTA) resulting from emotional *vs*. neutral stimuli confirmed that hippocampal gamma peaks locked to ongoing amygdala theta oscillations following all emotional stimuli (**Fig. 2E**, for one representative patient, and fig. S14 for remaining patients), in keeping with theta-gamma PAC results derived via the MI approach (**Fig. 2D**). The same relationship was observed between hippocampal theta and gamma (**Fig. 2F**, fig. S15). Critically, comparing PTAs for successful (eR) *vs*. unsuccessful encoded (eKF) aversive scenes revealed a theta phase difference between the two conditions (**Fig. 2G**), an effect observed across all patients (fig. S16).

The importance of this latter observation is that a potential function of phase-amplitude coupling in supporting memory formation is the facilitation of synaptic plasticity (*26*), which, in the case of theta-gamma PAC, is thought to depend on theta phase (*27*). In the context of the current findings, a phase-dependent mechanism underlying emotional memory formation suggests that amygdala theta- hippocampal gamma PAC is evoked by all emotional stimuli, but that the magnitude of the gamma amplitude is concentrated at different amygdala theta phase bins for subsequently remembered *vs*. not remembered emotional stimuli. We tested this explicitly, by calculating the Phase-Amplitude Coupling Opposition index (PACOi, Materials and Methods), which quantifies the strength of phase opposition between two different stimulus types (*i.e.*, eR *vs*. eKF; fig. S17). In line with the PTA results, hippocampal gamma amplitude during successful *vs*. unsuccessful encoding of aversive scenes concentrated at different theta bins with a consistent phase difference of ∼1.67 radians, corresponding to 30-45 ms (**Fig. 3A**). This was observed for all amygdala-hippocampus electrode pairs (fig. S18). This result established a phase code for amygdala theta-to-hippocampus gamma coupling in determining aversive memory formation in humans.

**Fig 3.**
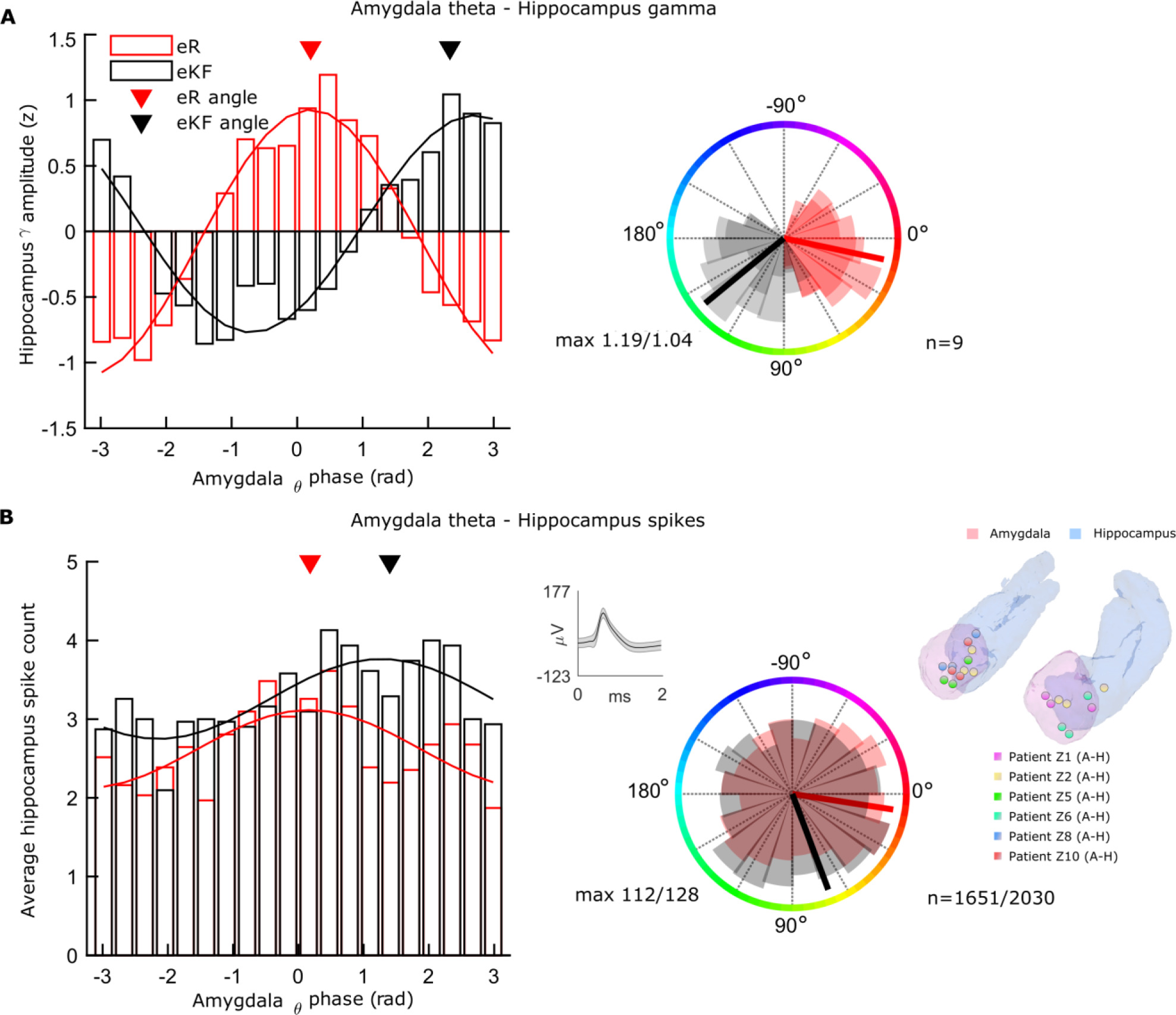
Emotional memory-dependent amygdala phase opposition of hippocampal gamma and single neuron activity. (**A**) Hippocampus gamma amplitude locks to different phase bins of amygdala theta depending on emotional memory outcome. Histogram of amygdala theta phases at which hippocampus broadband gamma activity occurs for eR (red) and eKF (black) trials as bar (left) and circular (right) plots. Inverted triangles represent the angle of preference (0.16 radians eR; 1.83 radians eKF) after phase realignment. Shaded areas in the circular plot represent PACOi results (*n*=9 electrodes) per phase bin (n=20) for each condition; maximum value of PAC for each condition is indicated (eR:1.19; eKF:1.04). **(B)** As for A but showing amygdala theta phases at which hippocampal spikes occur, average spike count over neurons *n*=31 neurons. Inverted triangles represent the angle of preference (0.18 radians eR; 1.40 radians eKF) after phase realignment. Shaded areas in the circular plot represent spikes per the 20 phase bins for each condition. Maximum spike count for each condition is indicated (eR: 112; eKF:128) as well as the total spike counts across neurons (eR: 1651, eKF:2030). Upper subpanel: single-neuron waveform (mean ± std) for one example neuron. Right side: Summary of electrode contact localization in the left and right amygdala (pink) and hippocampus (light blue) for patients included in the SFC analysis (Cohort 2: *n=*6 patients and *n*=7 electrodes; Patient Z2 had electrodes in bilateral hippocampus).

Our second cohort comprised patients with microelectrode recordings, permitting testing the derived phase-dependent mechanism at the single neuron level. That is, with Cohort 1 we considered gamma activity as an index of spiking activity (*23, 24*) and with Cohort 2 (*n*=6 patients, 7 electrodes) directly tested whether amygdala theta (3-12 Hz) organizes hippocampal neuronal firing by performing spike field coherence analysis (SFC) on simultaneous recordings of hippocampal single neurons and amygdala oscillatory activity (**Fig. 3B**, fig S1 B). Overall, we observed that hippocampal spikes (from *n*=31 neurons) during successful *vs*. unsuccessful encoding of aversive scenes concentrated at different theta bins with a consistent phase difference of ∼1.22 radians (two sample Kolmogorov- Smirnov test, *P*=0.00072), corresponding to ₥26 ms time lag. At the single-cell level, the firing distribution of 6 neurons differed significantly (circular Kuiper test, fig S21) between the two conditions, significantly more than expected by chance (binomial test versus 5% chance, *P*< 0.01) (**Fig. 3 B**).

Given that amygdala gamma power (**Fig. 1, D-E**) and phase-dependent amygdala theta-coupled hippocampal gamma activity (**Fig. 3A**) both tracked successful subsequent remember responses for emotional stimuli, an important question remained: are these two broadband gamma activities related? Although measures of coherence between amygdala and hippocampus did not show significant effects in the 50–150 Hz range, it remained possible that successful encoding of aversive scenes involved transient connectivity between broadband gamma activity bursts in both structures, to which standard measures of coherence would be insensitive. We therefore computed amygdala-hippocampus transient connectivity in the time range in which we observed significant amygdala high gamma activity power changes for successful encoding of aversive scenes (0.41–1.1 s; fig. S3). We selected windows of broadband gamma activity (60–120 Hz) as a proxy of neuronal activity(*23, 24*) around the hippocampal gamma peaks for both hippocampus and amygdala responses, and shifted these windows in time to compute the cross-correlogram for successful and unsuccessful encoding of aversive scenes. The shift in time is important because the observed phase opposition between PAC for two conditions (*i.e.*, eR *vs*. eKF) implies a condition-dependent phase lead/lag between the high frequency activity in hippocampus relative to the theta phase cycle in amygdala (**Fig. 4, A-B**). However, we do not know whether broadband gamma activity in amygdala shows a similar lead/lag time difference relative to hippocampal broadband gamma. Using the amplitude envelope as a measure of synchrony strength between the two structures, we indeed found stronger amygdala to hippocampus transient gamma synchronization for subsequently remembered compared to not remembered aversive scenes (**Fig. 4C**, fig. S19, all trials, lag 0.044–0.056 s, summed t value=29.73, *P=*0.011; controlling the number of trials between conditions, lag 0.044–0.06 s, summed t value=31.00, *P=*0.011).

**Fig. 4.**
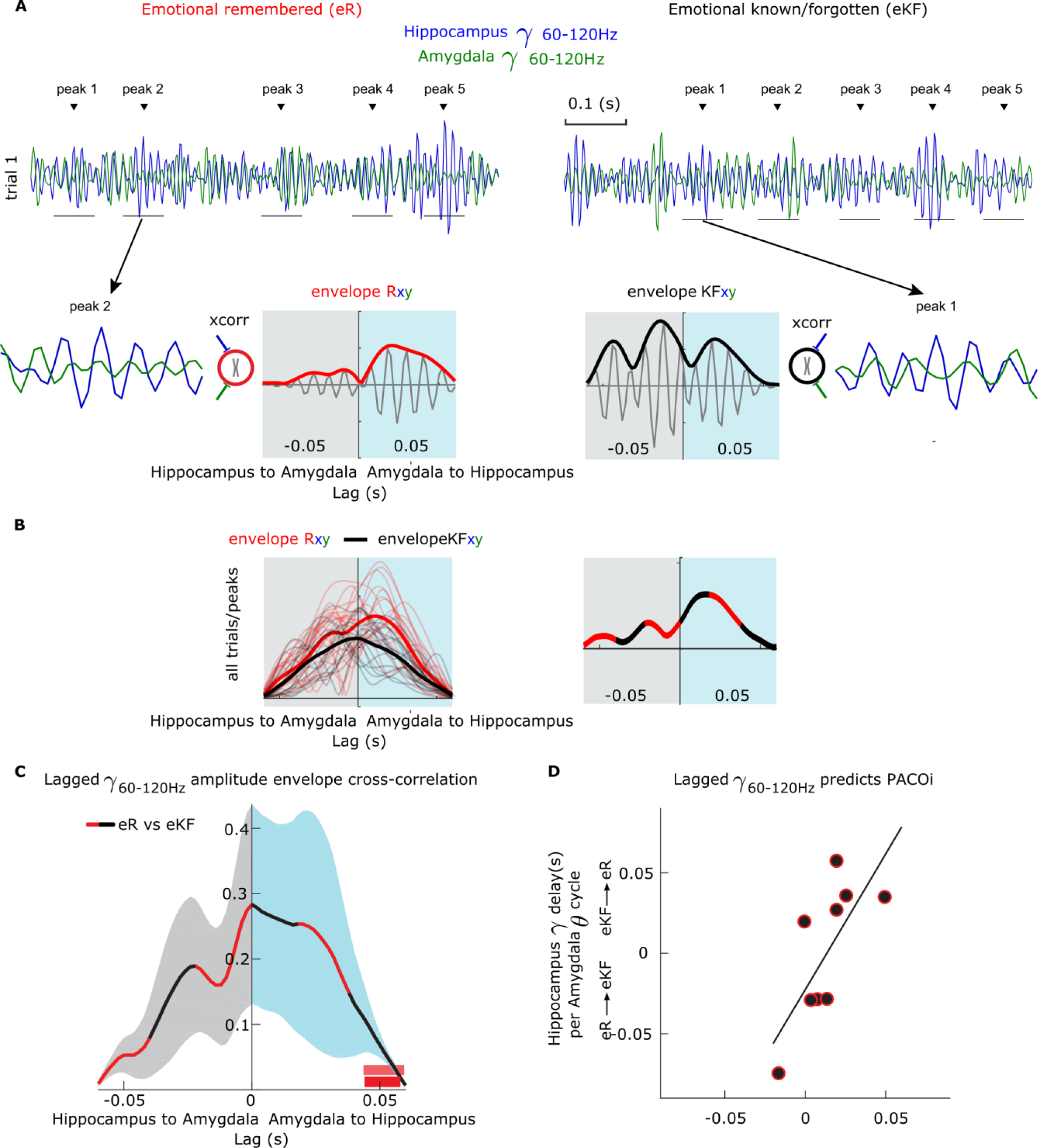
Broadband gamma (60–120 Hz) transient connectivity between amygdala and hippocampus is paced by amygdala theta-hippocampal gamma phase opposition. **(A-B)** Schematic depiction of the analysis. (**A**) Broadband gamma activity during aversive remembered (eR; red) trials was bandpass filtered (60–120 Hz) and peaks in hippocampal recordings were identified (top; data from two example trials from one patient). Short epochs around the identified peaks were cut (± 0.03 s; shown as thin horizontal lines under the gamma traces). For each epoch, the amplitude envelope of the cross-correlation between hippocampus (blue) and amygdala (green) epoch was computed for eR (below, red envelope) and eKF (black) trials. (**B**) The amplitude envelopes from all peaks and single trial cross-correlations (thin lines) were averaged and contrasted (averaged amplitude envelope eR - averaged amplitude envelope eKF; thick lines). The negative *x*-axis values of the envelope cross-correlogram indicate that hippocampus gamma leads amygdala gamma, whereas positive x-axis values indicate the reverse directionality. (**C**) Following the analysis in A and B in all patients in Cohort 1, amplitude envelope cross-correlation shows amygdala leading hippocampus broadband gamma activity. The shaded contours represent ± s.e.m and the color the directionality of the transient coupling. Red bar depicts significant lag window for all trials; light red bar above after controlling the number of trials between conditions. **(D)** Correlation between successful *vs*. unsuccessful emotional memory (eR-eKF) as measured as hippocampus-amygdala transient broadband gamma activity (fig. S18) peak lag and hippocampus gamma delay as a function of amygdala theta cycle. The averaged absolute value of *y*-axis values is 37.2 ± 5.9 ms.

To verify the role of ongoing amygdala theta in the modulation of the transient gamma synchronization between amygdala and hippocampus, we tested whether the transient connectivity indexed by the amplitude envelope and PACOi results were related. To this end, we converted the PACOi phase differences to time delays and correlated these estimates with the peak of the envelope of the cross-correlation computed as the difference between subsequently remembered *vs*. not remembered aversive trials (fig. S19). In line with the Granger causality results showing theta amygdala to hippocampus directionality effect, we found that amygdala gamma bursts lead hippocampal gamma bursts (Spearman rho=0.81, *P*=0.011; **Fig 4D**), with a latency of 37.2±5.9 ms (mean±s.e.m.). This significant correlation indicates a role of amygdala theta in synchronizing amygdala and hippocampal high gamma activity.

In sum, our data show that in response to an aversive visual stimulus, the amygdala influences ongoing hippocampal theta oscillations, which in turn organize the amplitude of local gamma activity and neuronal firing. Theta-gamma phase-amplitude coupling in the human hippocampus has been shown to be a general mechanism for memory encoding (*28, 29*). In rodents, theta-modulated gamma stimulation applied to BLA is an efficient protocol to enhance hippocampal CA1 gamma responses (*30*), and directly stimulating the human amygdala at theta-modulated gamma frequency leads to memory enhancement for simultaneously presented emotionally neutral stimuli (*31*). Here, we showed that emotional memory enhancement does not simply depend on theta-gamma phase-amplitude coupling. Confirming theoretical positions that memory formation is phase-dependent in humans (*19, 20*), we showed that the formation of memories for emotional stimuli (allowing later recollection) depends on the amygdala theta phase to which the hippocampal gamma and related neuronal firing couples. Critically, the phase difference associated with this process translates to the time lag required for amygdala and hippocampal gamma bursts to reach transient time-lagged coherence (∼25-45 ms). It is possible that this lag is related to the time required for noradrenergic input, upon which emotional memory formation is critically dependent (*12, 13*), to reach the medial temporal lobe, or that amygdala theta phase-dependent effects in the hippocampus are linked to the optimal conditions required for “emotion tagging” of memory (*32*) to occur. Moreover, these findings could represent a general mechanism through which amygdala oscillations influence other brain areas to enable emotion-induced modulation of further aspects of cognition, including perception, attention and decision-making (*14, 33*), and inform therapeutic approaches of amygdala stimulation to psychiatric disorders.

## Supplementary Materials

### Materials and Methods

#### Participants

##### Cohort1

Participants were 13 medication-resistant presurgical epilepsy patients with depth electrodes surgically implanted to aid seizure focus localization (Table S, fig S1A). Implantation sites were chosen solely on the basis of clinical criteria. Patients had normal or corrected-to-normal vision and had no history of head trauma or encephalitis. All patients had electrodes implanted in the amygdala and all amygdalae were radiologically normal on the pre-operative MRI.

We analyzed electrophysiological responses from 17 amygdalae from 13 patients (five had left, 4 right, and 4 bilateral medial temporal electrodes in the amygdala). Eleven patients also had electrodes in the ipsilateral anterior hippocampus. The hippocampal recording of one patient was excluded after hippocampal sclerosis was reported on the same side as the unilateral electrode implantation. For the second patient showing hippocampal sclerosis, only the non-pathological side was included. One further patient did not meet our criteria for spike-free trials (75%). We therefore analyzed electrophysiological responses from 9 hippocampi from 8 patients (1 patient had bilateral hippocampal electrodes). No statistical methods were used to pre-determine sample sizes but our sample sizes are larger or equal to those reported in previous publications (*16, 34*). Intracranial event-related potentials to the emotional and neutral pictures described, independent of subsequent memory, have been reported in 7 of the 13 patients presented here (*34*).

##### Cohort 2

This included 6 medication-resistant presurgical epilepsy patients implanted in the medial temporal lobe for diagnostic purposes. We analyzed neuronal signals from *n*=7 electrodes, simultaneously recording single neurons from the hippocampus and local field potentials (LFP) from the amygdala (Table S2, fig S1B). Note that patient Z1 is included in both cohorts; right medial temporal lobe in Cohort 1 and left in Cohort 2.

All patients signed informed consent. The study had full approval from the local ethics committees of the Hospital Ruber Internacional, Madrid, Spain and Kantonale Ethikkommission, Zurich, Switzerland (PB-2016-02055).

#### Stereotactic electrode implantation

For the patients recorded at the Hospital Ruber Internacional (Cohort 1), a contrast enhanced MRI was performed pre-operatively under stereotactic conditions to map vascular structures prior to electrode implantation and to calculate stereotactic coordinates for trajectories using the Neuroplan system (Integra Radionics). DIXI Medical Microdeep depth electrodes (multi-contact, semi rigid, diameter of 0.8 mm, contact length of 2 mm, inter-contact isolator length of 1.5 mm) were implanted based on the stereotactic Leksell method. For the patient recorded in Zurich, Switzerland (Cohort 2 and patient Z1 in Cohort 1), the depth electrodes (1.3 mm diameter, 8 contacts of 1.6 mm length, and spacing between contact centers 5 mm; Ad-Tech, Racine, WI, www.adtechmedical.com) were stereotactically implanted into the amygdala, hippocampus, and entorhinal cortex. Each macroelectrode had nine microelectrodes that protruded approximately 4 mm from its tip.

#### Electrode contact localization

To localize electrodes, we used the procedure described previously (*34*). For each patient, the post-electrode placement CTs (post-CT) was co-registered to the pre-electrode placement T1- weighted magnetic resonance images (pre-MRI). To optimize co-registration, both brain images were first skull-stripped. For CTs this was done by filtering out all voxels with signal intensities between 100 and 1300 HU. Skull stripping of the pre-MRI proceeded by first spatially normalizing the image to MNI space employing the New Segment algorithm in SPM8 (http://www.fil.ion.ucl.ac.uk/spm). The resultant inverse normalization parameters were then applied to the brain mask supplied in SPM8 to transform the brain mask into the native space of the pre-MRI. All voxels in pre-MRI lying outside the brain mask and possessing a signal value in the highest 15th percentile were filtered out. The skull-stripped pre-MRI was then co-registered and re-sliced to the skull-stripped post-CT. Next, the pre-MRI was affine normalized to the post-CT, thus transforming the pre-MRI image into native post-CT space. The two images were then overlaid, with the post-CT thresholded such that only electrode contacts were visible. Electrode contacts for each patient are shown in fig. S1 A-B in native space.

##### Electrode contact visualization

Skull-stripped pre-MRI and post-CT were normalized to MNI space, as described in (*35*). The three-dimensional view of amygdala and hippocampus is an average of the segmented bilateral amygdala and hippocampus from each patient’s pre-operative MRI (segmentations done using FreeSurfer v.6 software, https://surfer.nmr.mgh.harvard.edu/). Electrode contact locations for both Cohorts patients are displayed using Paraview (www.paraview.org) in Fig. 1C and 3B. In the few cases where patients had electrodes implanted in both anterior and posterior hippocampus, only the anterior electrode contacts were included in our analyses, given the greater anterior *vs*. posterior hippocampal connectivity with amygdala (*6*).

#### Experiment

##### Stimuli

Patients were presented with 40 emotional and 80 neutral color pictures during the encoding session. These were drawn at random from a pool of 80 high-arousing aversive (mutilations and attack) scenes selected from the International Affective Picture System (IAPS)(*36*), and 160 low-arousing neutral pictures: 149 taken from the IAPS (household scenes and neutral persons) and eleven neutral landscape pictures taken from the world-wide web. Mean normative IAPS picture ratings (s.e.m.) on a nine-point scale for valence were 5.05 (± 0.05) for neutral, and 2.04 (± 0.05) for aversive pictures, and for arousal were 3.29 (±0.06), and 6.3 (±0.07) for neutral and aversive pictures, respectively. Emotional items are better remembered than neutral ones (*8*), thus the ratio of aversive to neutral stimuli was 1:2 to promote a balanced number of trials per condition (Table S5).

##### Procedure

Prior to signing informed consent, patients were shown one example of an aversive IAPS picture and instructed that they would see similar pictures both on that day and the next. Task instructions were provided both verbally and on-screen in Spanish (Cohort 1) and in German for patients recorded in Switzerland (Cohort 2). Encoding and recognition sessions were conducted during the third and fourth post-operative days, respectively, in Madrid, and second and third post- operative days in Zurich (Fig. 1A). During the encoding session, emotional and neutral pictures were presented pseudo-randomly (presentation time 0.5 s; interstimulus interval 3.5 s) with a constraint that emotional pictures were separated by at least one neutral picture. Pictures were displayed on a 27x20.3 cm video monitor (1024x768 pixels) placed at a distance of ⁓50 cm from the subject’s eyes (30.2° x 22.9°). Patients were required to make an indoor-outdoor judgment to each picture via button-press. On average they made a button press to 99.07% ± 0.30% (mean ± s.e.m) of the trials. After 24 hours, patients were presented with all stimuli from the encoding session, randomly intermixed with 120 foils (40 emotional and 80 neutral pictures). Patients were required to make a “remember”, “know”, “new” decision (R-K-N) (*37*). During both sessions, patients remained as still as possible attending the centre of the screen while avoiding verbalisations and minimising eye- blinks.

#### Data Acquisition

At the Ruber Hospital Internacional, Madrid (Cohort 1), ongoing intracranial EEG (iEEG) activity was acquired using an XLTEK EMU128FS amplifier (XLTEK, Oakville, Ontario, Canada). Intracranial EEG data were recorded at each electrode contact site at a 500 Hz sampling rate (online bandpass filter 0.1–150 Hz) and referenced to linked mastoid electrodes. Three patients were recorded at 2000 Hz sampling rate but data were down-sampled at 500 Hz. Intracranial data in Zurich (Cohort 2) were acquired with a Neuralynx ATLAS system with sampling rate 4000 Hz (online band-pass filter of 0.5–1000 Hz) and recorded against a common intracranial reference and then down-sampled at 500 Hz.

#### Data analysis

##### Pre-processing

Intracranial EEG data analysis was performed using the FieldTrip toolbox (https://www.fieldtriptoolbox.org) (*38*) running on Matlab version R2017b (the Mathworks, Natick, MA, USA). For all patients, recordings were transformed to a bipolar derivation by subtracting signal from adjacent electrode contacts within the hippocampus or amygdala. Previous studies demonstrate that bipolar referencing optimizes estimates of local activity (*39–41*) and connectivity patterns between brain regions (*42*). For each amygdala and hippocampus bipolar channel, experimental condition, and patient, epochs from -7.5–7.5 s peri-stimulus time with respect to picture onset were extracted from continuous iEEG data. Epochs were then de-trended and no off-line filtering was applied. Trials containing signal artifacts caused by epileptic spikes or electrical noise were detected on visual inspection in the time domain and removed (percentage of trials without epileptic spikes or noise for each patient is reported in Table S5). Note, that when patients had electrodes implanted in the amygdala and the hippocampus, trials were rejected if an artefact was detected in either or both structures (Table S6).

##### Spectral analysis

Time-resolved spectral decomposition was computed for each trial using 7 Slepian multi-tapers for high frequencies (> 35 Hz) and a single Hanning taper for low frequencies (≤35 Hz). The selected Slepian tapers for the analysis of high frequencies were based on windows of 0.4 s width and a 10 Hz frequency smoothing. The time-resolved spectral estimation was done in 2.5 Hz steps. In contrast to the high frequency analysis where we used a constant time-frequency smoothing, sliding windows were defined by 7 cycles per frequency step for the low frequency analysis. Trial-by-trial visual inspection and artefact rejection was then repeated in the time-frequency domain. Trials with interictal epileptiform activity (such as fast high frequency activity followed by lower frequency power) as well as excessive noise, including broadband noise from hospital equipment were removed (*43*).

Time-frequency estimates were then baseline corrected by calculating the relative percentage change with respect to baseline (-1 to -0.1 s pre-stimulus time). The spectral activity was then averaged over channels within each structure (amygdala and hippocampus) and trials for each patient.

##### Statistics

To test for subsequent memory effects, we specified six effects of interests. The event at encoding corresponding to aversive and neutral pictures separated according to whether they were later remembered, or either received a familiarity judgement or were forgotten (eR, eKF, nR, nKF). Given the low number of aversive forgotten trials compared to aversive remember ones, we merged know and forgotten trials for both emotional and neutral ones and performed the analysis on this set of data. Note, that repeating the spectral power analysis (described below) comparing remember *vs.* forgotten trials showed analogous frequency effect to the power analysis in which we compared remember *vs.* know trials (fig. S4–S5). We focused our analyses on encoding responses to subsequently remembered *vs*. known/forgotten items. In both amygdala and hippocampus, we first tested for the interaction of emotion by successful encoding (aversive remember (eR) – aversive known/forgotten (eKF)) *vs.* (neutral remember (nR) – neutral known/forgotten (nKF)). Power interactions were computed using 8 bipolar channels obtained from 7 patients. One patient was excluded (Patient Z1) because only 2 trials in the neutral remembered condition were obtained. In absence of a significant interaction, we tested for the main effect of emotion (aversive – neutral), and the main effect of successful encoding (remember – known/forgotten).

To correct the family wise error rate in the context of multiple comparisons across time and frequency dimensions, we applied a cluster-based permutation test (*44*) to the ensuing *t-*values obtained at each time-frequency bin to determine significant interactions or main effects in the time-frequency domain. At each permutation step, selecting the a-priori time of interest in our data (from 0–1.5 s), clusters were formed by temporal and frequency adjacency with cluster threshold of *P=*0.05. Permutation steps were repeated 10000 times. After each permutation step, a paired *t*-test was calculated at each time and each frequency bin for high (35–150 Hz) and low frequencies (0–34 Hz) separately using a threshold of p=0.025. For every dependent measure (time-frequency power, connectivity measures, etc) we only report the significant results unless explicitly stated.

To control for potential pre-stimulus power effects during the baseline period, we performed cluster-based permutation test during the baseline time window to test for main effects of memory, and emotion, and their interaction, for amygdala and hippocampus recordings (n=8). None of the contrasts yielded significant condition differences in the baseline period.

##### Connectivity analyses

For all connectivity analyses, only the most lateral electrode contact pairs (bipolar channels) in the amygdala and the hippocampus were included. Given that the depth of electrode implantation (*i.e.*, how medial the electrode tip is placed) varies across patients, taking the most lateral contact pairs promotes homogeneity of contact localization within the same lateral portion of the amygdala and hippocampus sampled across patients. By doing so, it is likely that electrode contacts included in our analyses are placed within (baso)lateral amygdala and CA1 of the hippocampus. Note that we repeated the time-frequency power analysis using the most lateral channels (fig. S5-S6) and we found analogous frequency effects (fig. S6).

##### Coherence

To investigate interactions between regions, we first computed spectral coherence using Fieldtrip. We subtracted the event-related potential from each single trial and computed a fast Fourier transform (FFT) using the single taper von Hann method with 7 cycles per time window (-0.2 to 1.5 s in 0.1 s steps) from 2–150 Hz. Entering the magnitude of coherence for the same frequency range, we tested for main effects of emotion, successful encoding and the emotion by memory interaction (eR-eKF vs. nR-nKF) followed by non-parametric cluster-based permutation correction for multiple comparisons (see above).

##### Time and frequency resolved Granger causality

This analysis was computed using the Fieldtrip and BSMART (http://www.brain-smart.org/”) toolboxes (*45*). To ensure covariance stationarity (*46*), the mean-corrected time series was submitted to a Kwiatkowski-Phillips-Schmidt-Shin (KPSS) test (*47*) for each brain region (amygdala and hippocampus), subject, and condition and it was stationary. The time domain data was first low-pass filtered at 85 Hz and then down-sampled to 250 Hz. We computed time-dependent sets of multivariate autoregressive coefficients in overlapping windows of 0.4 s length (moving forward in 1-time point steps from -0.5 to 1.5 s) (*45*). The optimal model order was estimated for each patient, most lateral electrode pair and condition by means of the Bayesian Information criterion (BIC). We selected a model order of 9 so to capture Granger causality for all subjects within a sufficiently large time window (corresponding to a time lag of 0.036 s). The order of Granger causality corresponded to a lag of 36 ms, which is in the range of latencies observed for human hippocampal responses to direct electrical stimulation of the amygdala (10-40 ms) (*48*). Thereafter, Granger causality (2-34 hz) was calculated based on the transfer matrices computed from the autoregressive coefficients. In order to assess statistically the directionality between the amygdala and the hippocampus, the Granger coefficients were compared using a cluster-based permutation test to quantify main effects of emotion, successful encoding and the interaction between them (eR-eKF vs. nR-nKF) in both directions (amygdala to hippocampus and hippocampus to amygdala), as described previously (*16, 49*).

##### Phase to amplitude coupling: Modulation index (MI) and PACOi

As unequal trial numbers can confound phase estimates, we controlled for this as follows. For each patient, contact and experimental condition, the lowest number of trials was determined (N_min_). From the other condition, a subset of Nmin observations was randomly drawn from the observations constituting that particular experimental condition and the index of interest was computed. This procedure was re-computed 20 times by randomly selecting the subset of trials to the rest of the experimental conditions with higher number of trials. The controlled subsampled index was the average taken over the subsampled estimates.

##### Phase to amplitude modulation index

We calculated phase-to-amplitude coupling (PAC) using the Modulation index (MI) as previously defined (*25, 50*). This was performed over the time period where we found a subsequent memory by emotion interaction within amygdala gamma broadband activity (0.41–1.1 s; Fig. 1H). First, for each patient and bipolar channel of interest, single trials were bandpass filtered around two sets of frequencies to obtain instantaneous phase (<u>f_p_</u>; from 5–20 Hz in steps of 1 Hz) and amplitude (<u>f_a_</u>; from 40–120 Hz in steps of 5 Hz). The analytic phase (<u>φf_p_</u>) was obtained by taking the angle of the Hilbert transform of the bandpass filtered data around <u>f_p_</u> with a bandwidth of ± 2 Hz. For the same trial, the analytic amplitude (a<u>f_a_</u>) was similarly obtained but by taking the magnitude of the Hilbert transform of the bandpass filtered data around fa with a bandwidth of ± 15 Hz. We used FIR filters with the filter order being set to three cycles of the lower bandwidth bound. Before filtering, single trials were z-scored. Second, for a given frequency pair, we constructed the amplitude-phase histograms as follows. The analytic phase signal was divided in *n*=20 equal bins (<u>φ_n_=</u>[-π π]) and the mean analytic amplitude was taken over those specific bins. Third, for each frequency pair, the MI is computed as the Kullback-Leibler divergence (*51*) between the amplitude-phase histogram pooled from all corresponding trials and compared to the uniform distribution.

##### Phase-Amplitude Coupling Opposition index (PACOi)

The MI measures the modulation of the amplitude of high frequencies by the phase of low frequencies independently of the preferred phase angle at which the high frequency amplitude occurs. We derived a novel metric, the PACOi, to exploit potential differences in the preferred phase angle that two PAC distributions can produce. For example, the maximum at which the amplitude of an oscillation (*i.e.*, gamma) concentrates in a given phase bin (*i.e.*, theta peak) in each experimental condition can be different (or not) from another condition (*i.e.*, theta trough). To formalize this quantification, we tested whether the pair-wise phase consistency (PPC) of each trial type exceeded the overall PPC taken over the two groups of trials together (*52*). Fig. S17 provides, schematically, the rationale and quantification of the measure. The calculation of PACOi starts by first transforming the amplitude-phase histograms (see Modulation index above) to the complex domain as follows:

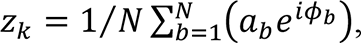

with *k* being a single trial of a particular experimental condition, *N* the total number of phase bins, and *ab* and *ϕ*_*b*_ representing the amplitude and the angle of a specific bin *b*. Therefore, we obtain a complex number per single trial and experimental condition. Once in the complex domain, these indices are normalized to unit length (*z*/abs(*z*)) and the PPC is computed for each condition separately (PPC1, PPC2), and then by combining the trials of the two conditions (PP_Call_; fig. S17). This procedure was carried out, with the rationale that if the gamma amplitude in eR and eKF concentrates systematically at different bins of theta, then the sum of the PPC obtained from each experimental condition separately should be larger than the PPC obtained from all trials pooled. Statistical significance was computed by randomly shuffling (n=1000) the trial condition assignment (remembered *vs.* known/forgotten) (*52*). It is a common observation that the angular bin to which the phase opposition points varies between test subjects (*52, 53*). Each patient’s phase distributions were therefore realigned (Fig. 3A) such that, for each patient, the amygdala theta phase at which the hippocampus broadband gamma coupled was set to a phase angle of zero under the eR condition. The exact number of phase shifts was obtained by computing the angle at which the vector strength was pointing at (angle(⟨ z_eR_ ⟩), being ⟨ ⟩ the average over complex single trials). Once the angle was obtained (*i.e.*, π/2) this angle was subtracted from both the eR and eKF conditions. Thus, eR theta-gamma PAC histogram is centered around zero. However, eKF angle preference can fall either (-π 0] or [0 π), which is a nontrivial property confirming our hypothesis that eR and eKF are associated with opposite phase angles. By convention, we mirror flipped the eKF histogram distribution when the vector strength angle falls with the (-π 0].

##### Inter-region peak-triggered averages (PTA)

We computed PTA as a complementary way to quantify the intra- and inter-regional PAC. First, we band-pass filtered the gamma band from a particular brain region (amygdala/hippocampus) using a two-pass finite impulse response (FIR) filter with a filter order of 3 cycles in the lowest frequency bound. Peaks were detected (0.1 s minimum inter-peak distance) in this filtered signal and time intervals of ± 0.12 s around these peaks were used to average the raw traces (z-scored) over the same or different brain regions. Once averaged, the PTAs were de-trended. Cross-correlations between the averaged PTAs of two conditions served to compare PTAs from two conditions (Fig. 2 E-G and fig. S14–S16). Power spectral density (PSD) of the cross-correlated PTAs was computed to find the main spectral component that dominates the PTA (PSD peak; f_peak_). Best sinusoidal fit was computed over the cross-correlated PTA with frequency, phase, amplitude and offset as free parameters. The frequency parameter was constrained selecting the frequency range of ± f_peak_/4.

##### Broadband gamma transient connectivity analysis

This analysis measured transient connectivity (envelope cross-correlations) between amygdala and hippocampal broadband gamma activity. First, we band-pass filtered the time series from the amygdala and the hippocampus contacts between 60–120 Hz (*23, 24*) using a two-pass finite impulse response (FIR) filter with a filter order of 3 cycles of the lower bound. This range was chosen because it encompasses both amygdala and hippocampal gamma ranges. Before bandpass filtering, single trials were demeaned. Hippocampal gamma peaks were detected (0.1 s minimum inter-peak distance) in this filtered signal and ± 0.03 s epochs were selected. For each epoch, the amplitude envelope of the single-trial cross-correlation between hippocampus and amygdala was computed (see Fig. 4 A-B). The single-trial amplitude envelope was computed by taking the magnitude of the Hilbert transform. Finally, single trial amplitude envelopes were averaged for each contact and experimental condition.

##### Relationship between within vs. cross-frequency hippocampus-amygdala coupling

The correlation reported in Fig. 4D was computed as follows. For each patient, the peak of the amplitude envelope of the cross-correlation for eR minus eKF was taken. This comparison shows both the magnitude and direction of the transient broadband gamma activity between hippocampus and amygdala relative to each experimental condition. On the other hand, for each patient and bipolar channel, PACOi was computed under the eR *vs*. eKF contrast. The resulting significant phase differences found between the high frequency hippocampus amplitude as a function of amygdala low frequency (theta) phase for eR *vs*. eKF were transformed to the time domain as follows:

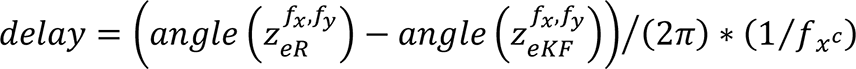

with *f_x_c* being the center of mass taken over the amygdala low frequencies, *z* being the PACOi, *f_x_* and *f_y_* representing the x (gamma) and y (theta) significant PACOi pairs of frequencies (fig. S17). Delay was the output time index used in the correlation, representing the time difference (*i.e.*, lead/lag) between hippocampus high frequency amplitude in eR and eKF given a low frequency amygdala phase. To test the strength of the we performed a surrogate analysis in which we permuted the patients delay and peak envelope association and computed the Spearman rho after each shuffling (n=10000). The observed rho was then compared to the permutation distribution.

##### Spike detection and sorting

In 6 patients with microelectrodes placed in the hippocampus (Cohort 2, *n*=7 electrodes, one patient had bilateral hippocampal electrodes, fig S1B), we recorded broadband neuronal activity (Neuralynx, sampling rate 32 kHz), which we separated into single- or multi neuron activity and LFP. Neuronal spikes were detected and sorted using Wave_Clus (*54*). We refer here to a putative neuron by the term “neuron.” The default settings were employed with the following exceptions: “template_sdnum” was set to 1.5 to assign unsorted spikes to clusters in a more conservative manner; “min_clus” was set to 60 and “max_clus” was set to 10 in order to avoid over-clustering; and “mintemp” was set to 0.05 to avoid under-clustering. All clusters were visually inspected and judged based on the spike shape and its variance, inter-spike interval (ISI) distribution, and the presence of a plausible refractory period (*55*). If necessary, clusters were manually adjusted or excluded.

For recording quality assessment (fig. S20), we calculated (1) the number of neurons recorded on each wire, (2) the ISI refractoriness for each neuron, (3) the mean firing rate for each neuron, and (4) the waveform peak signal-to-noise ratio (SNR) for each neuron. The ISI refractoriness was assessed as the percentage of ISIs with a duration of <3ms. The waveform peak SNR was determined as: SNR=A_peak_/STD_noise_, where A_peak_ is the absolute amplitude of the peak of the mean waveform, and STDnoise is the standard deviation of the raw trace (filtered between 300 and 3000 Hz). We isolated zero, one or two distinct neurons from each microelectrode. In every trial, the number of spikes occurring between 0.41 and 1.1 seconds was calculated and spikes with ISIs < 3ms were removed. We included *n*= 33 neurons in the hippocampus for further analysis.

##### Spike field coherence

We examined the relationship between the hippocampal neuronal action potentials and the amygdala LFP oscillations. The LFP recordings were taken from the macroelectrode recording from the ipsilateral amygdala, except for two patients in whom the LFP was extracted from an ipsilateral amygdala microelectrode due to a system data acquisition error. Macroelectrode data were recorded at 4 kHz. We down-sampled recordings to 2 kHZ as previously reported(*56*) and then applied a 40 Hz low pass filter. To calculate the amygdala theta phase we filtered the data from 3 to 12 Hz. The analytic phase was obtained by taking the angle of the Hilbert transform of the bandpass filtered data for each trial in the condition eR and eKF. The analytic phase signal was divided in n=20 equal bins (φ_n_=[-π π]) and for each condition separately we extracted the amygdala theta phase at which each hippocampal spike occurred and calculated the number of spikes occurring at each bin in each condition. For each neuron and each condition, we averaged over bins (using circ_mean in the Circular Statistics Toolbox (*57*)) and obtained the preferential phase in radians to depict spike theta coherence. Since the angular bin to which the SFC points varies between subjects we realigned each patient’s phase distributions as explained above (see PACOi method).

Thus, we obtained a spike-field coherence histogram for each neuron. For each neuron we performed a circular Kuiper test and compared eR vs eKF distributions (fig S21). We further tested whether the number of significant neurons was above chance using a binomial test. For the group level analysis, a two-sample Kolmogorov-Smirnov test on the average results from all observed neurons (*n*=31) was performed (Fig 3B).

## Data Availability

All data needed to evaluate the conclusions in the paper are present in the paper and/or the
supplementary materials. Data and code will be made publicly available after publication

---

**Fig. S1.**
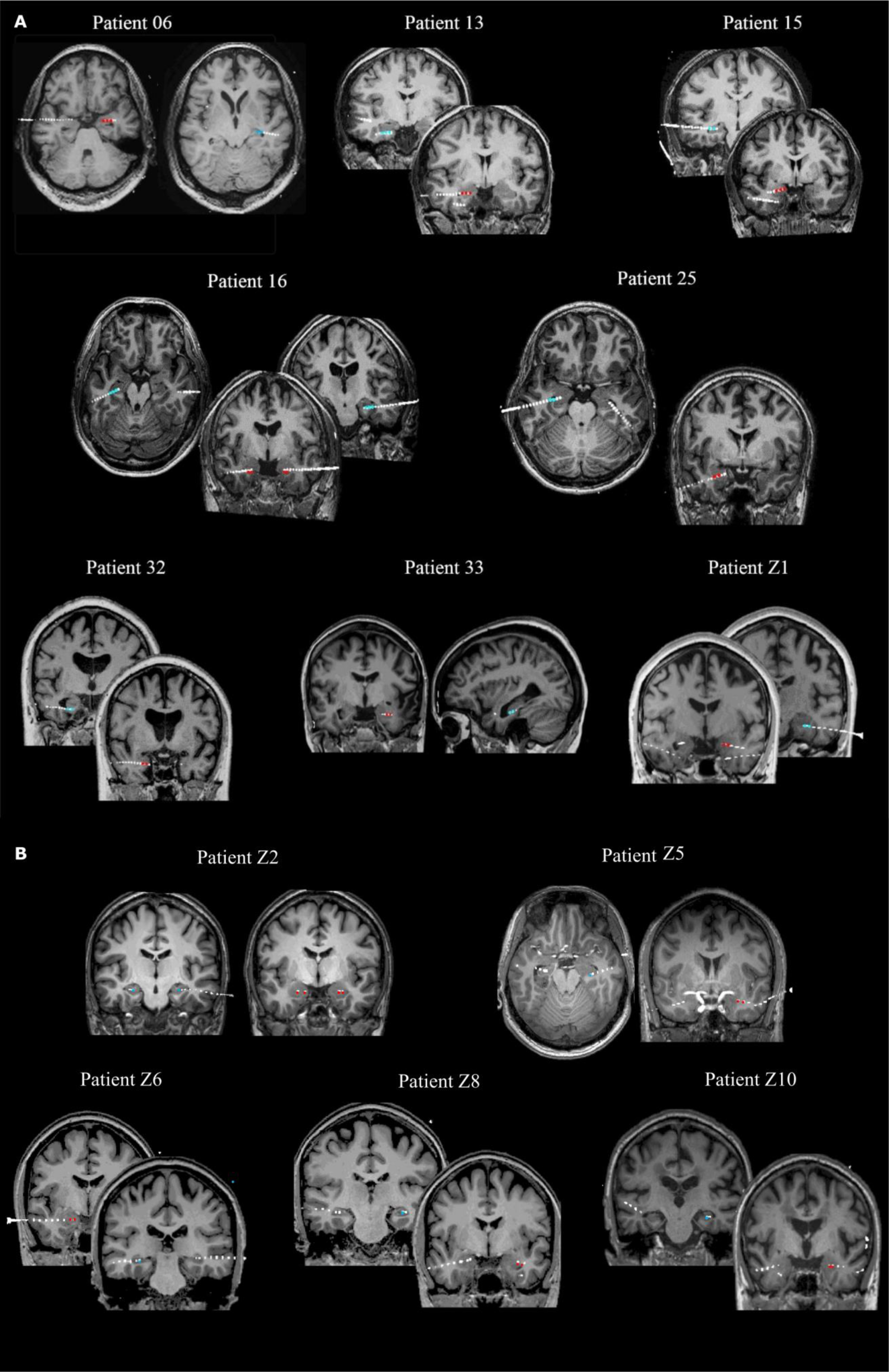
(**A**) **Electrode contact localization for the 8 patients (Cohort 1) with macro-electrodes localized in both amygdala and hippocampus.** Coronal sections are shown for all patients; sagittal view is shown for patient 33, and axial sections for patient 6, 16 and 25. Red dots indicate amygdala contacts and blues dots hippocampal contacts included in our analyses. (**B**) **Electrode contact localization for patients (Cohort 2) with macro-electrodes localized in both amygdala and hippocampus.** Coronal sections are shown for all patients; and axial sections for patient Z5, for better visualization of hippocampal contacts. Red dots indicate amygdala contacts. The blue dot represents the first hippocampal contact and point at putative microelectrode location. For all patients, post-operative CT images from each patient have been co-registered with their corresponding pre-operative MRI scans in native space and superimposed to display amygdala and hippocampal contacts (CTs have been thresholded so as to only show electrode contacts).

**Fig. S2.**
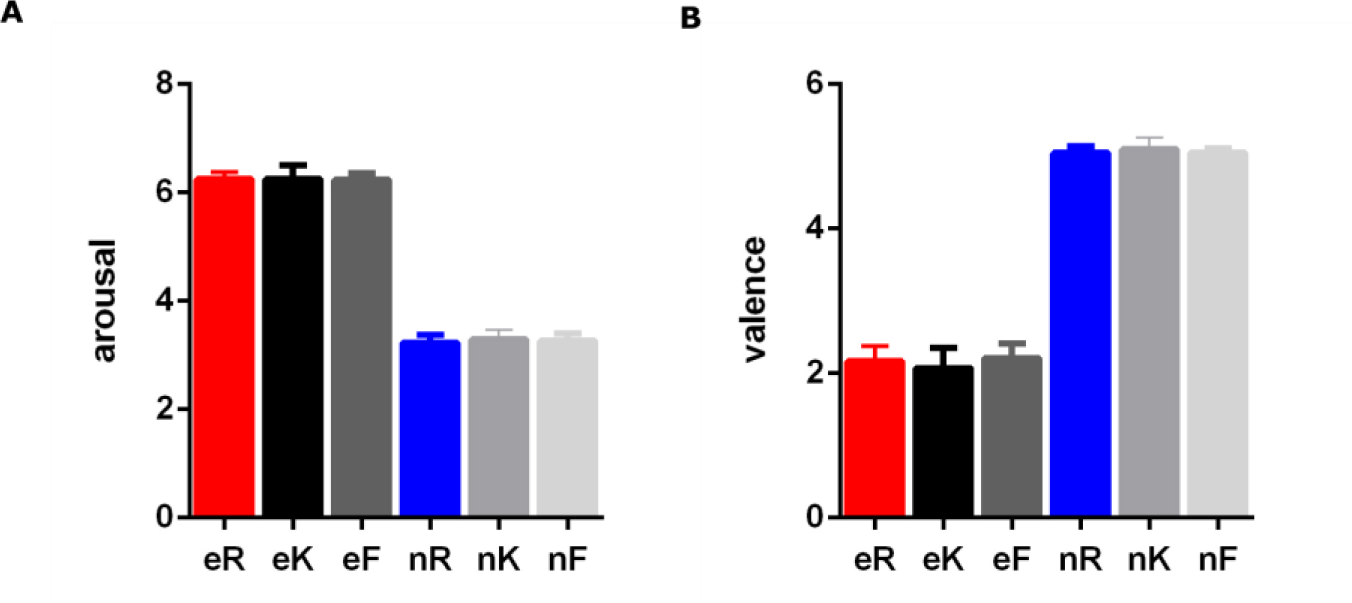
Emotional memory performance: subsequent remembering *vs*. knowing *vs*. forgetting is not related to differences in arousal and valence ratings. Aversive and neutral IAPS pictures were selected on the basis of normative ratings for arousal and valence. To test whether amygdala and hippocampal subsequent memory-dependent spectral responses to emotional scenes could have been driven by different arousal or valence, we performed the following comparisons. For all stimuli presented at encoding, normative arousal ratings were entered into a repeated measures ANOVA with factors emotion (aversive e, neutral n) and subsequent memory (R, K, F) for both patient cohorts 1 and 2 (*n*=18 patients). We did not observe a significant emotion by memory interaction (*F*_(2, 16)_ =1.93, *P=*0.169, η²=0.199). Repeating this comparison with valence ratings, we again do not show a significant interaction, (*F*_(2, 16)_ = 2.76, *P=*0.093, η²=0.25). There was no main effect of subsequent memory for either arousal (*F*_(2, 16)_=0.906, *P=*0.424 η²=0.102) or valence rating (*F*_(2, 16)_ =3.47, *P=*0.056, η²=0.303). The main effect of emotion is significant for both rating parameters, as stimuli were selected on this basis.

**Fig S3.**
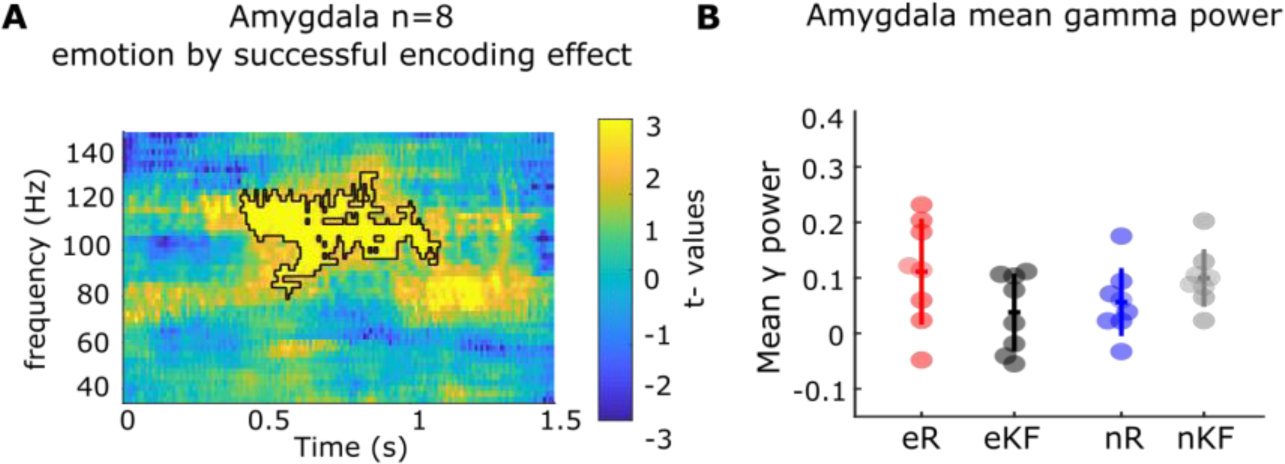
Amygdala gamma power during successful emotional memory formation in the group of patients with electrodes in both the amygdala and ipsilateral hippocampus (Cohort 1) (**A**) Time frequency resolved test statistics for the emotion by subsequent memory effect, (emotion by subsequent memory interaction, summed *t* value=1898.01, *P=*0.0042). Amygdala power change in the high gamma range is greater for eR *vs*. eKF relative to nR vs. nKF scenes. The colorbar indicates t-values (*Post-hoc t* tests on mean power changes show eR *vs.* eKF t_7_=2.40, *P=*0.047, d=0.85; nR *vs.* nKF, t_7_=-2.94, *P*=0.021, d=-1.04). (**B**) Mean amygdala gamma power in the significant cluster (from 0.41–1.1 s and from 80–132 Hz) for the four trials types. Circles show individual electrode data. Horizontal / vertical lines indicate mean / ± s.e.m. (*n*= 7 patients and 8 electrodes, one patient (Patient Z1) was excluded from power interaction testing as only 2 trials in the neutral remembered condition were obtained).

**Fig. S4.**
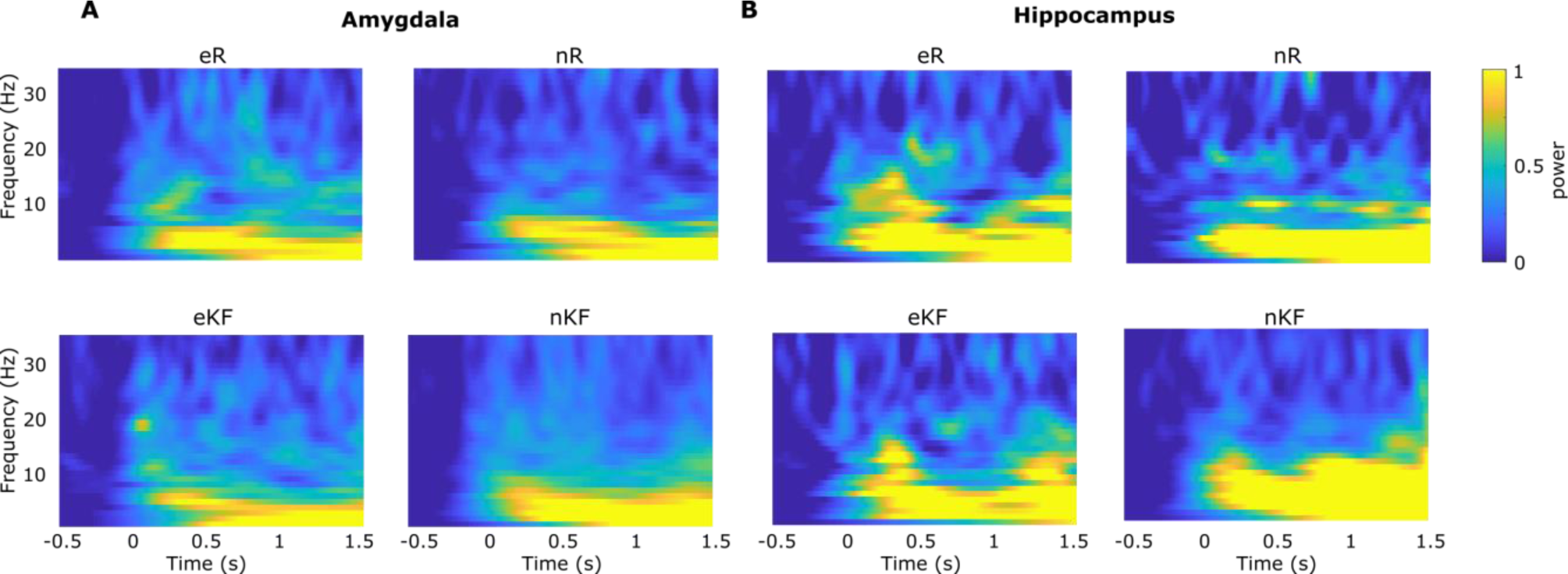
Stimulus-induced low frequency oscillatory power during encoding in amygdala and hippocampus (Cohort 1). Low frequency oscillatory relative power increases, primarily in the delta and theta ranges, are observed to all conditions in both (**A**) Amygdala and (**B**) Hippocampus. In neither structure did we observe an emotion by subsequent memory interaction, or main effect. eR: subsequently remembered and eKF know/forgotten aversive pictures; nR subsequently neutral remembered and nKF known/forgotten neutral trials. The colorbar indicates power change relative to baseline.

**Fig. S5.**
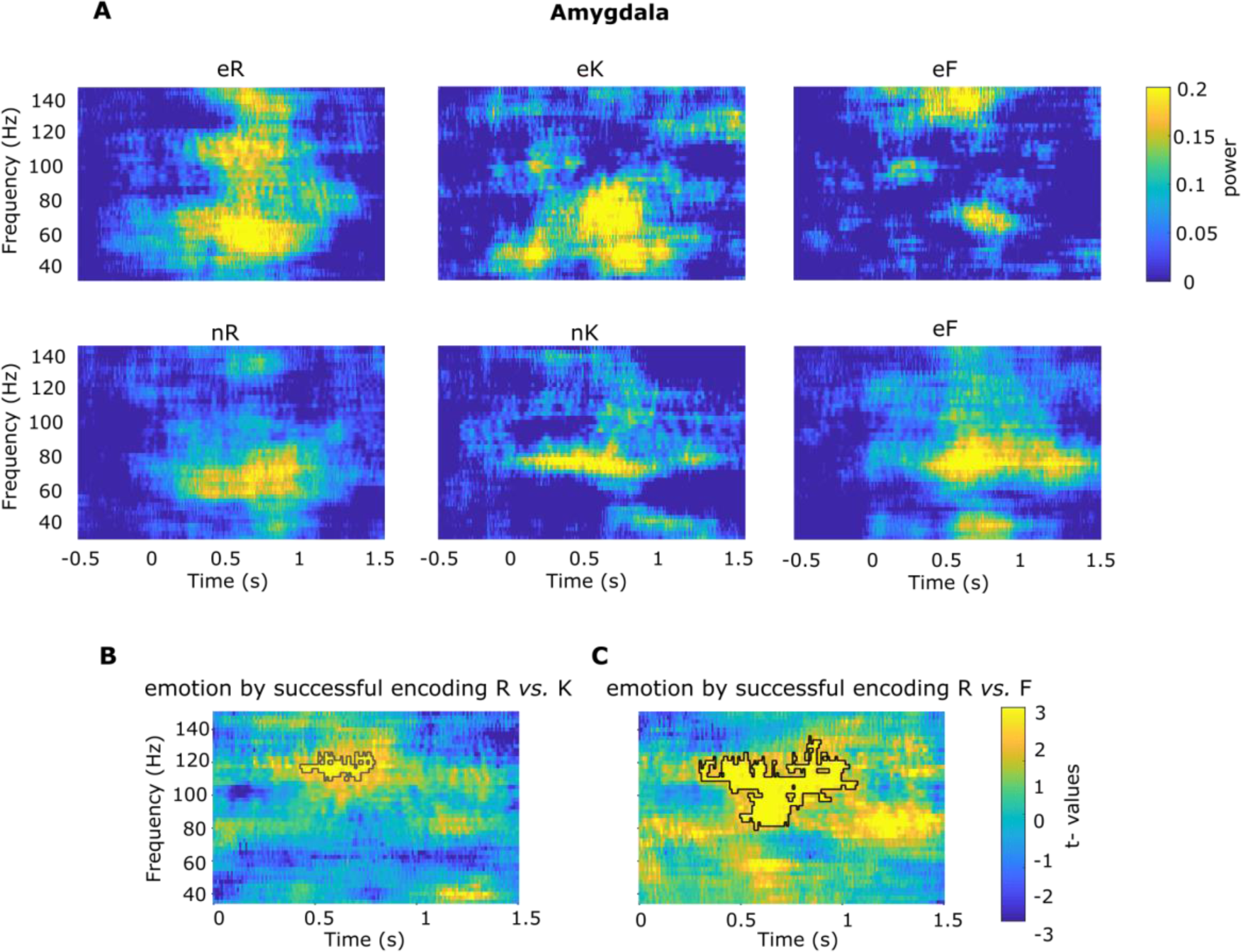
Fast amygdala gamma power during successful emotional memory formation: overlapping frequency effects when comparing gamma responses during aversive subsequent remember *vs*. both know (K) and forgotten (F) trials (Cohort 1). **(A)** Amygdala power change in the gamma range for the six trial types: emotional remember (eR), know (eK), forgotten (eF); neutral remember (nR), know (nK), forgotten (nF). Colorbar depicts relative change from baseline. **(B, C)** Time frequency resolved test statistics showing overlapping frequency effects when **(B)** testing the interaction (eR-eK) *vs*. (nR-nK) (summed t value=438.52, *P*=0.11, cluster from 0.43–0.79 s and from 110-125 Hz) (*n*=7 patients; 8 electrodes) and **(C)** the interaction (eR-eF) *vs*. (nR-nF) (summed t value=2306.6, *P*=0.01, cluster from 0.30–1.07 s and from 80-135 Hz), (*n*=7 patients; 8 electrodes). No significant gamma power effects (*P* > 0.05) were observed on comparing eK *vs*. eF trials. Colorbar depicts the t-statistic.

**Fig. S6.**
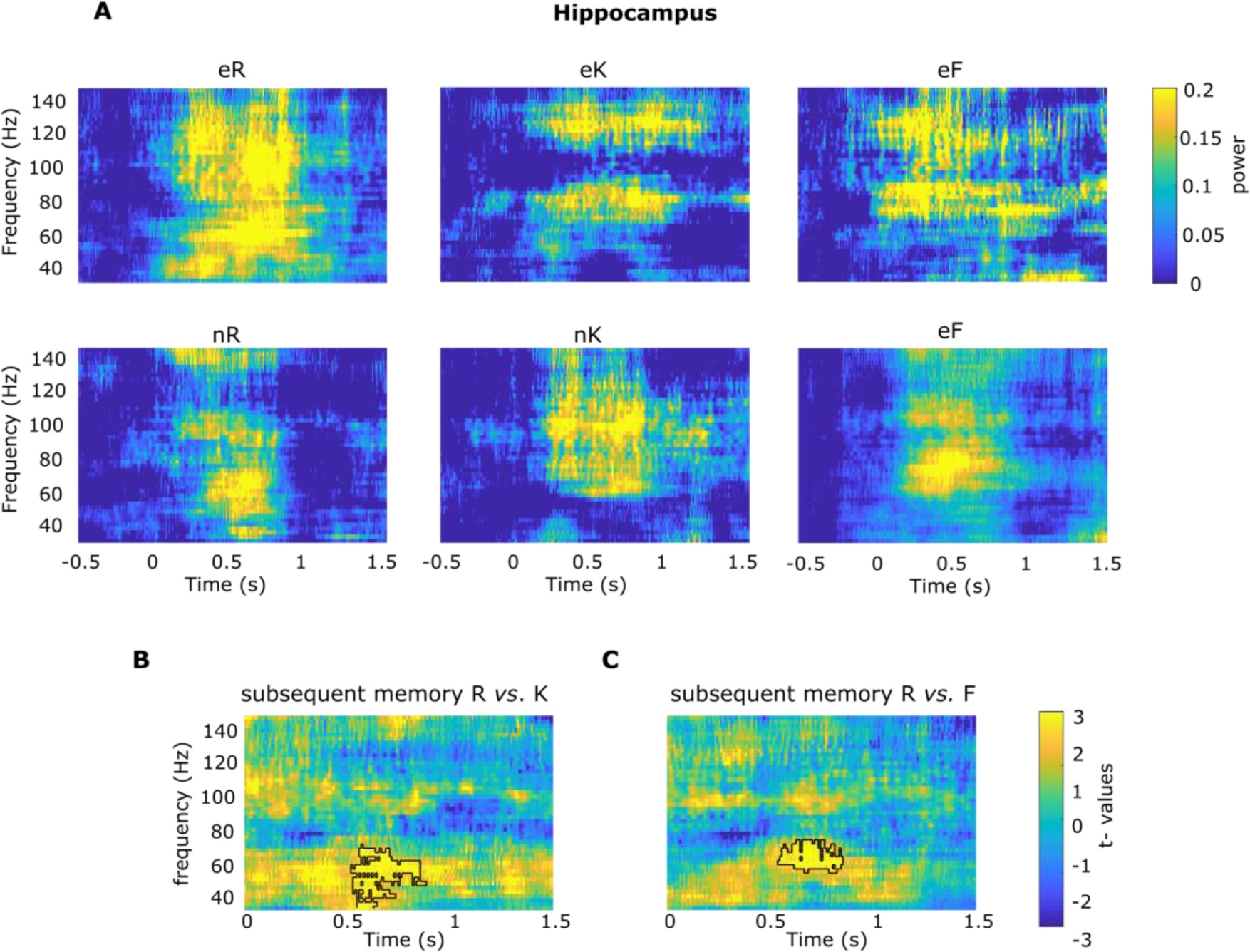
Gamma power in the hippocampus during memory formation: overlapping frequency effects when comparing gamma responses during subsequently remembered *vs*. both know (K) and forgotten (F) trials (Cohort 1). **(A)** Hippocampus power change in the gamma range for the six trial types: aversive remember (eR), know (eK), forgotten (eF); neutral remember (nR), know (nK), forgotten (nF). Colorbar depicts relative change from baseline. **(B, C)** Time frequency resolved test statistics showing overlapping frequency effects when comparing gamma responses during **(B)** subsequently remembered minus subsequently know (R-K) (summed t value=778.15, *P*=0.016, cluster from 0.52– 0.88 s and from 35-72 Hz) and **(C)** subsequently remembered minus subsequently forgotten (R-F) trials (summed t value=521.47, *P*=0.024, cluster from 0.54– 0.85 s and from 57-75 Hz), (*n*=8 patients; 9 electrodes). No significant gamma power effects (*P* > 0.05) were observed on comparing K *vs*. F trials in the hippocampus. Colorbar depicts the *t*-statistic.

**Fig S7.**
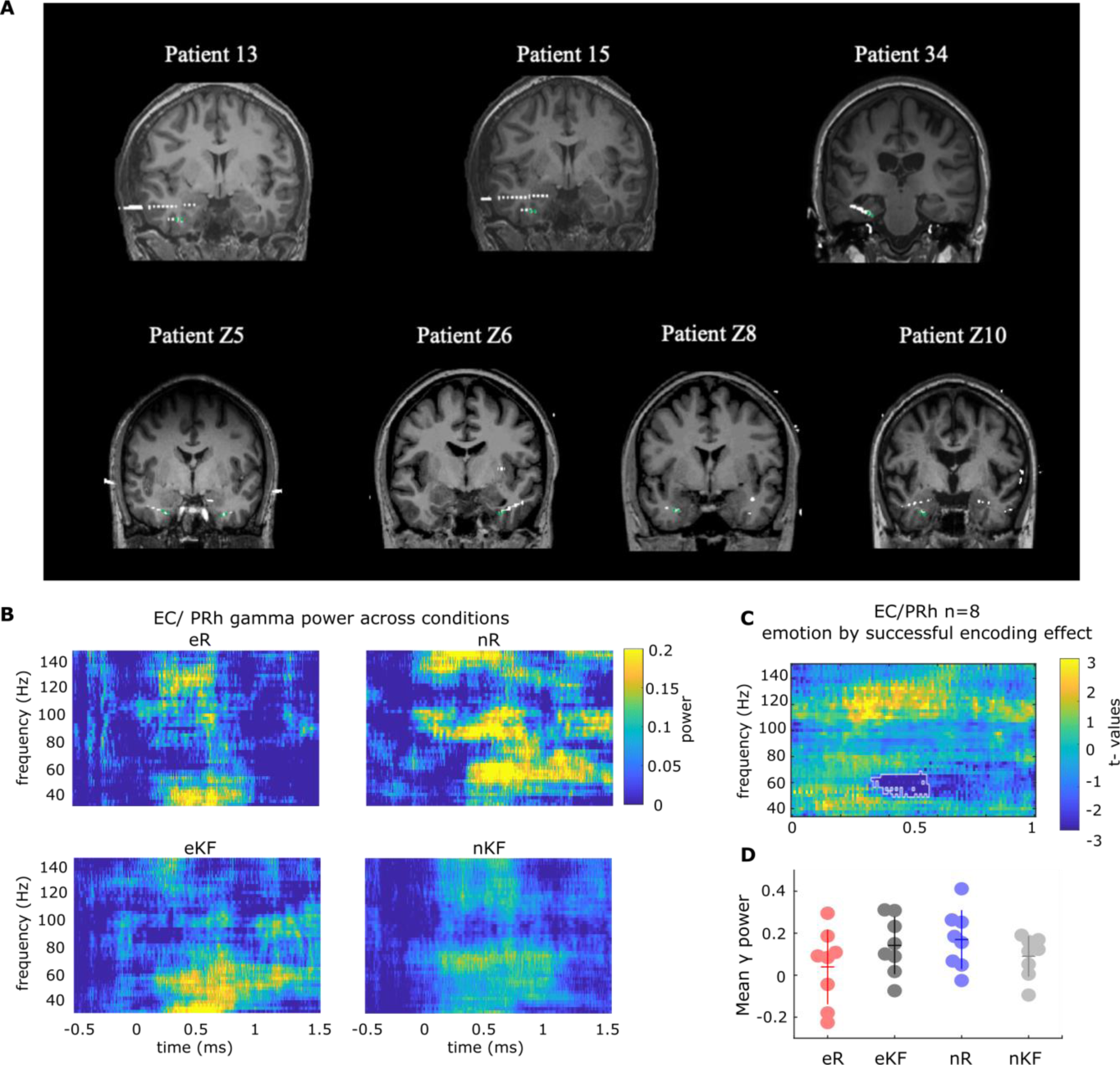
Entorhinal (EC) and Perirhinal (PRh) cortex gamma activity during memory formation of neutral, but not emotional, scenes. EC/PRh recordings were pooled over Cohorts 1 and 2 (*n*=8 electrodes) **(A) Electrode contact localization for the 7 patients with macro-electrodes localized in the Entorhinal (EC) or Perirhinal (PRh) cortex. Green dots indicate contacts included in the analyses. (B)** EC/PRh power change in the gamma range for the four trial types: emotional remember (eR), known/forgotten (eKF); neutral remember (nR), know/forgotten (nKF). Colorbar depicts relative change from baseline. (**C**) Time frequency resolved test statistics for the emotion by subsequent memory effect. EC/PRh power change in the gamma range is greater for nR *vs*. nKF relative to eR vs. eKF scenes, (emotion by subsequent memory interaction, summed *t* value=-475.27, *P*=0.023). The colorbar indicates t-values. (eR *vs.* eKF t_7_= -2.94, *P=*0.021; nR *vs.* nKF t_7_= 3.10, *P=*0.017) (**D**) Mean gamma power in the significant cluster (from 0.33 –0.56 s and from 50–67 Hz) for the four trials types. Circles show individual electrode data. Horizontal / vertical lines indicate mean / ± s.e.m.

**Fig. S8.**
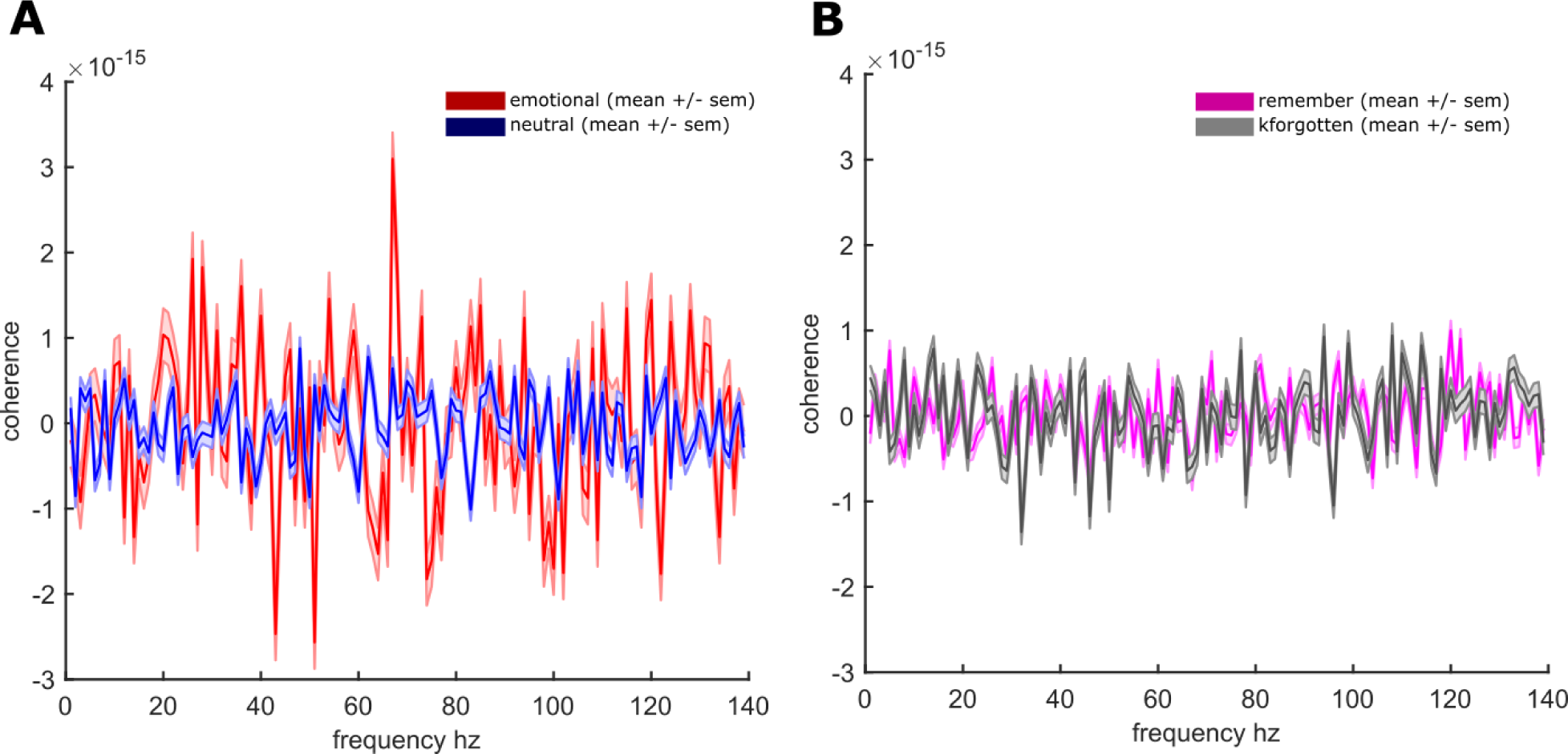
Coherence between stimulus-induced amygdala and hippocampal activity. Frequency-resolved coherence (mean ± standard error) between amygdala and hippocampus (Cohort 1, *n*= 8 patients; 9 amygdala-hippocampal electrode pairs) is plotted during **(A)** emotional *vs*. neutral trials, and **(B)** remember *vs.* know and forgotten trials. No main effects in any frequency were found for either comparison, nor did coherence values show an emotion (aversive, neutral) by subsequent memory (R, KF) interaction.

**Fig. S9.**
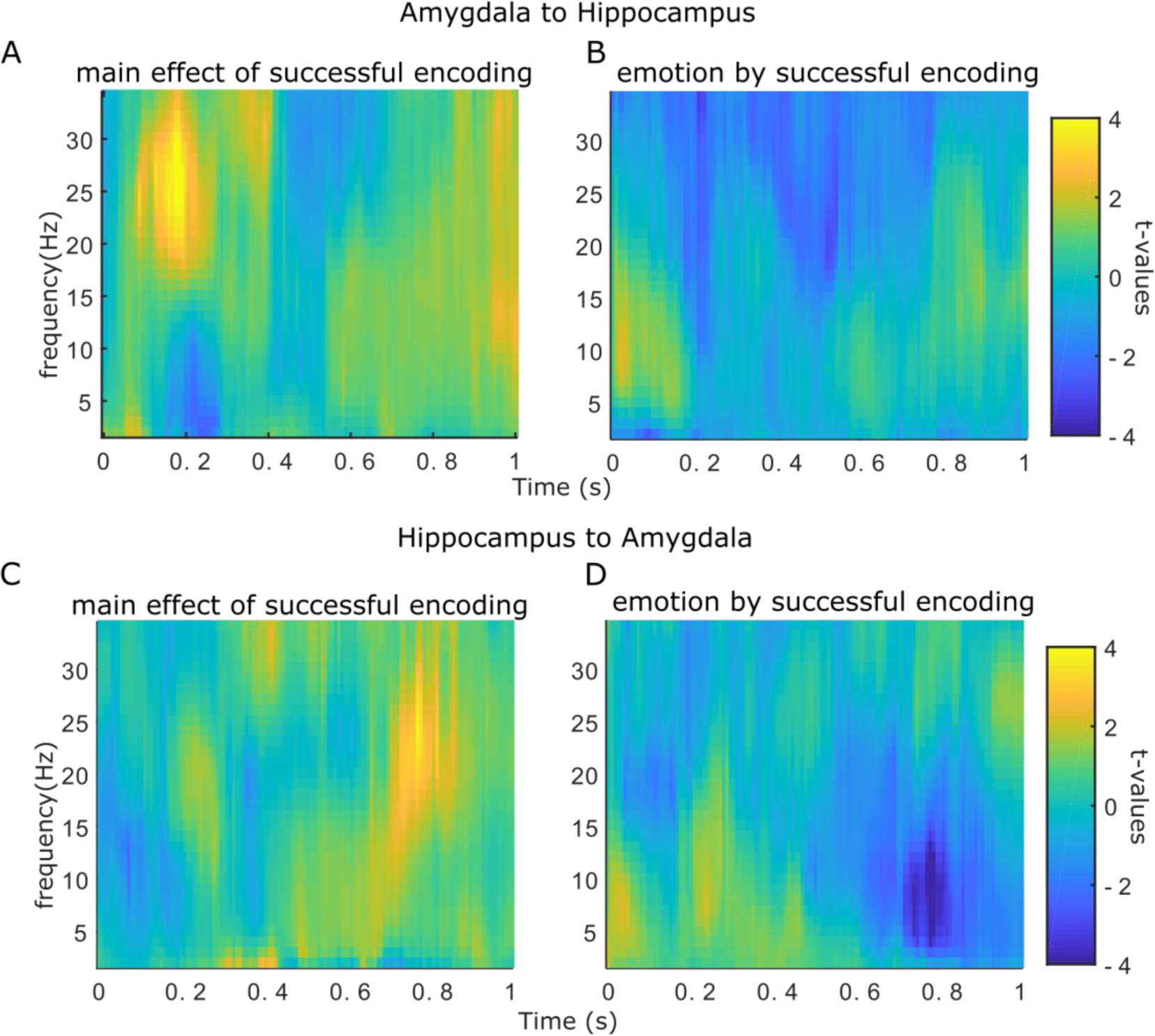
Time and frequency-resolved Granger causality (GC) between amygdala and hippocampal activity (Cohort 1). Time and frequency resolved GC in the amygdala to hippocampus direction is plotted for (**A**) the main effect of successful encoding (Remember vs Known/Forgotten and (**B**) the interaction emotion by successful encoding (eR-eKF) vs. (nR - nKF). The same measures, but in the hippocampus to amygdala direction, are plotted below for (**C**) the main effect of successful encoding and (**D**) the interaction emotion by successful encoding. No effect survived cluster-based correction.

**Fig. S10.**
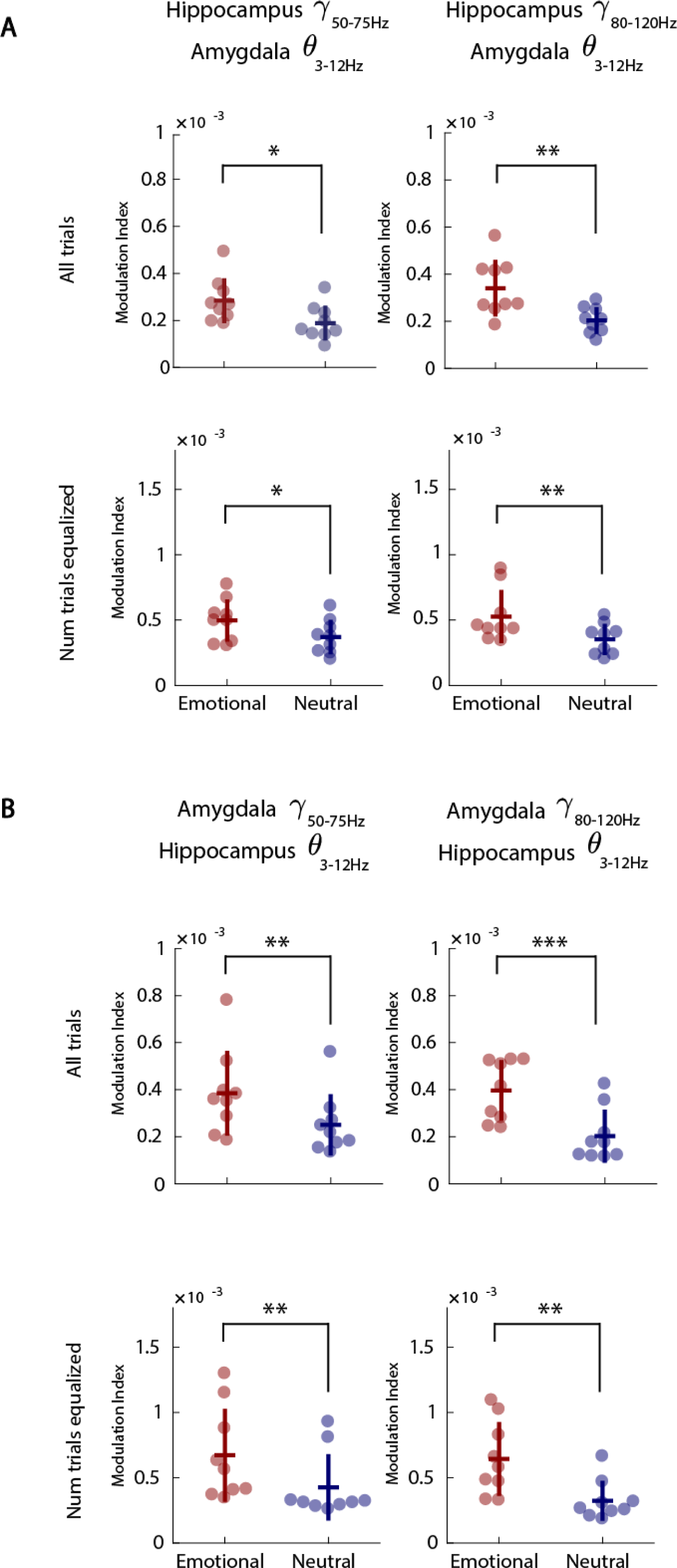
Between-region theta-gamma phase-to-amplitude coupling (Cohort 1). (**A**) Modulation index for PAC between amygdala theta phase (3–12 Hz) and hippocampus gamma amplitude, in the 50–75 Hz range (upper left, identical to Fig. 2D in the main text) and hippocampus gamma amplitude in the 80–120 Hz range (main effect of emotion: F_(1, 8)_=22.9, *P*=0.0014, η^2^=0.741). Lower left and right panels show the same conditions but controlling for the number of trials between conditions (F_(1, 8)_=6.65, *P=*0.033, η^2^=0.454; F_(1, 8)_=12.6, *P=*0.0075, η^2^=0.612, respectively; see Materials and Methods). For each dot plot, the modulation index for the emotion condition (eR+eKF) is shown in red and the neutral condition (nR+nKF) in blue. Horizontal and vertical bars represent the mean and s.e.m., respectively. Each dot represents a single bipolar channel in a given experimental condition. (**B**) Modulation index for PAC between hippocampus theta phase (3–12 Hz) and amygdala gamma amplitude, with same figure conventions as in (A). Upper left, hippocampus theta phase to amygdala gamma 50–75 Hz amplitude (main effect of emotion: F_(1, 8)_=20.3, *P=*0.0020, η^2^=0.718); and upper right, amygdala gamma 80-120 Hz amplitude (main effect of emotion: F_(1, 8)_=44.2, *P=*0.00016, η^2^=0.847). Lower left and right show the emotion main effect controlling the number of trials between conditions (amygdala gamma 50–75 Hz amplitude, F_(1, 8)_=15.1, *P=*0.0046, η^2^=0.654; amygdala gamma 80–120 Hz amplitude, F_(1, 8)_=19.7, *P=*0.0022, η^2^=0.711). All reported main effects were computed including lateral bipolar channels from all patients with simultaneous hippocampus and amygdala recordings (*n=*8 patients, 9 amygdala-hippocampal electrode pairs) during 0.41–1.1 s post-stimulus onset.

**Fig. S11.**
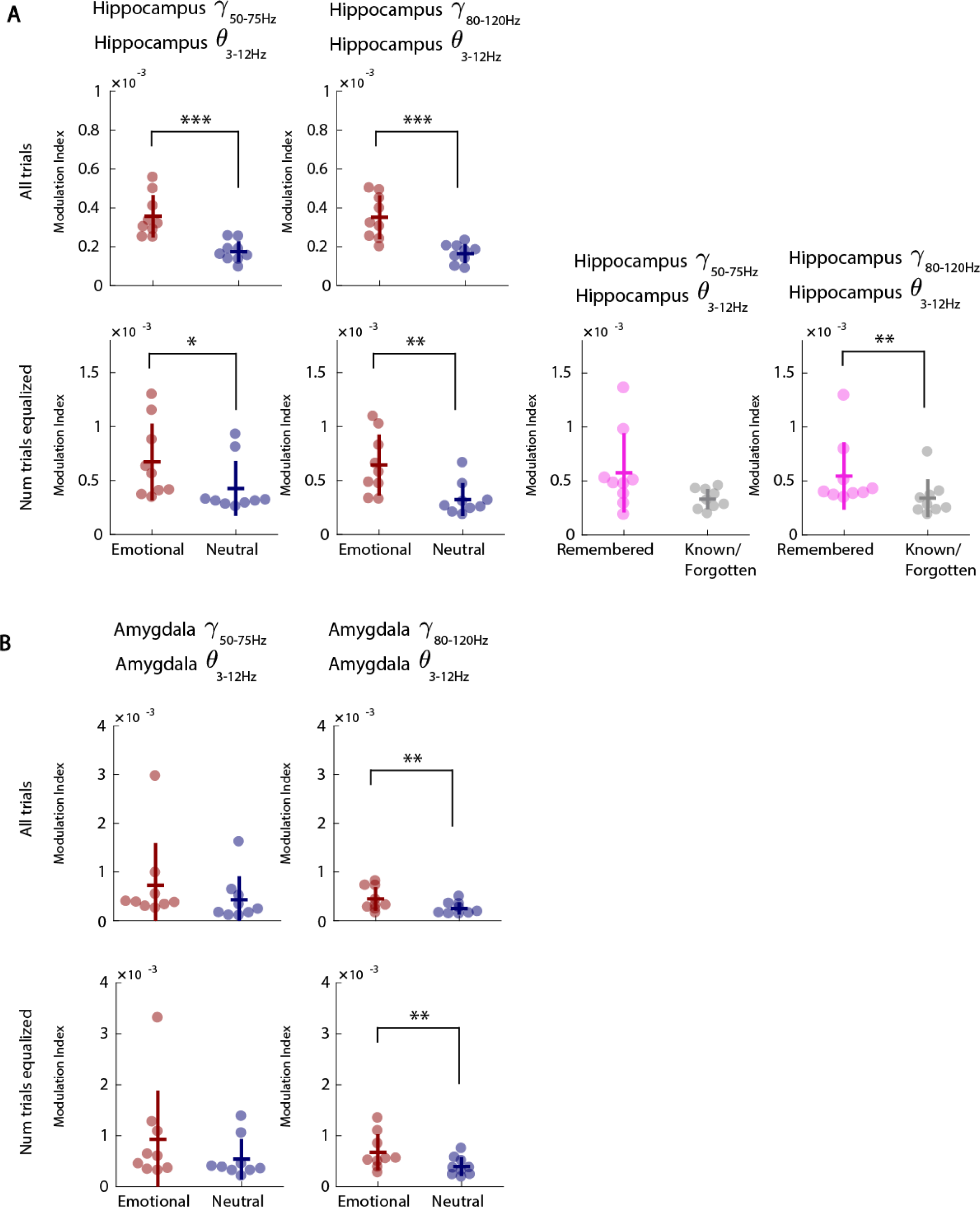
Within-region theta-gamma phase-to-amplitude coupling (Cohort 1). (**A**) Modulation index for PAC between hippocampus theta phase (3–12 Hz) to hippocampus gamma amplitude. Upper left: hippocampus gamma amplitude 50–75 Hz; emotion main effect F_(1, 8)_=27.5, *P*=0.00078, η^2^=0.775; Upper right: hippocampus gamma amplitude 80–120 Hz; emotion main effect F_(1, 8)_=42.8, *P*=0.00018, η^2^=0.843. Lower two left panels show the same conditions but controlling for the number of trials between conditions (F_(1, 8)_=10, *P*=0.013, η^2^=0.556; F_(1, 8)_=16.6, *P*=0.0036, η^2^=0.675, respectively). For each dot plot, the modulation index for the emotion condition (eR+eKF) is shown in red and the neutral condition (nR+nKF) in blue. Horizontal and vertical bars represent the mean and s.e.m., respectively. Each dot represents a single bipolar channel in a given experimental condition. The bottom right two dot plots show the modulation index for hippocampal theta phase to hippocampal gamma as a function of subsequent memory, with number of trials equalized between subsequently remembered (eR+nR, magenta) and not remembered (eKF+nKF, grey) trials. The main effect of memory was not significant for the hippocampus 50–75 Hz gamma band (F_(1, 8)_=5.08, *P*=0.054, η^2^=0.389) but it was evident for the hippocampus 80–120 Hz band (F_(1, 8)_=16.6, *P*=0.0036, η^2^=0. 675). (**B**) Modulation index for PAC between amygdala theta phase (3–12 Hz) to amygdala gamma amplitude. Upper left: amygdala gamma amplitude 50–75 Hz; emotion main effect F_(1, 8)_=4.07, *P*=0.08, η^2^=0.337; Upper right: hippocampus gamma amplitude 80–120 Hz; emotion main effect F_(1, 8)_=14.4, *P=*0.0052, η^2^=0.664. Lower two left panels show the same conditions but controlling for the number of trials between conditions (F_(1, 8)_=2.69, *P=*0.14, η^2^=0.252; F_(1, 8)_=12.2, *P=*0.0082, η^2^=0.603, respectively). All reported main effects were computed including lateral bipolar channels from all patients with simultaneous hippocampus and amygdala recordings (*n* =8 patients, 9 amygdala-hippocampal electrode pairs) during 0.41–1.1 s post-stimulus onset.

**Fig. S12.**
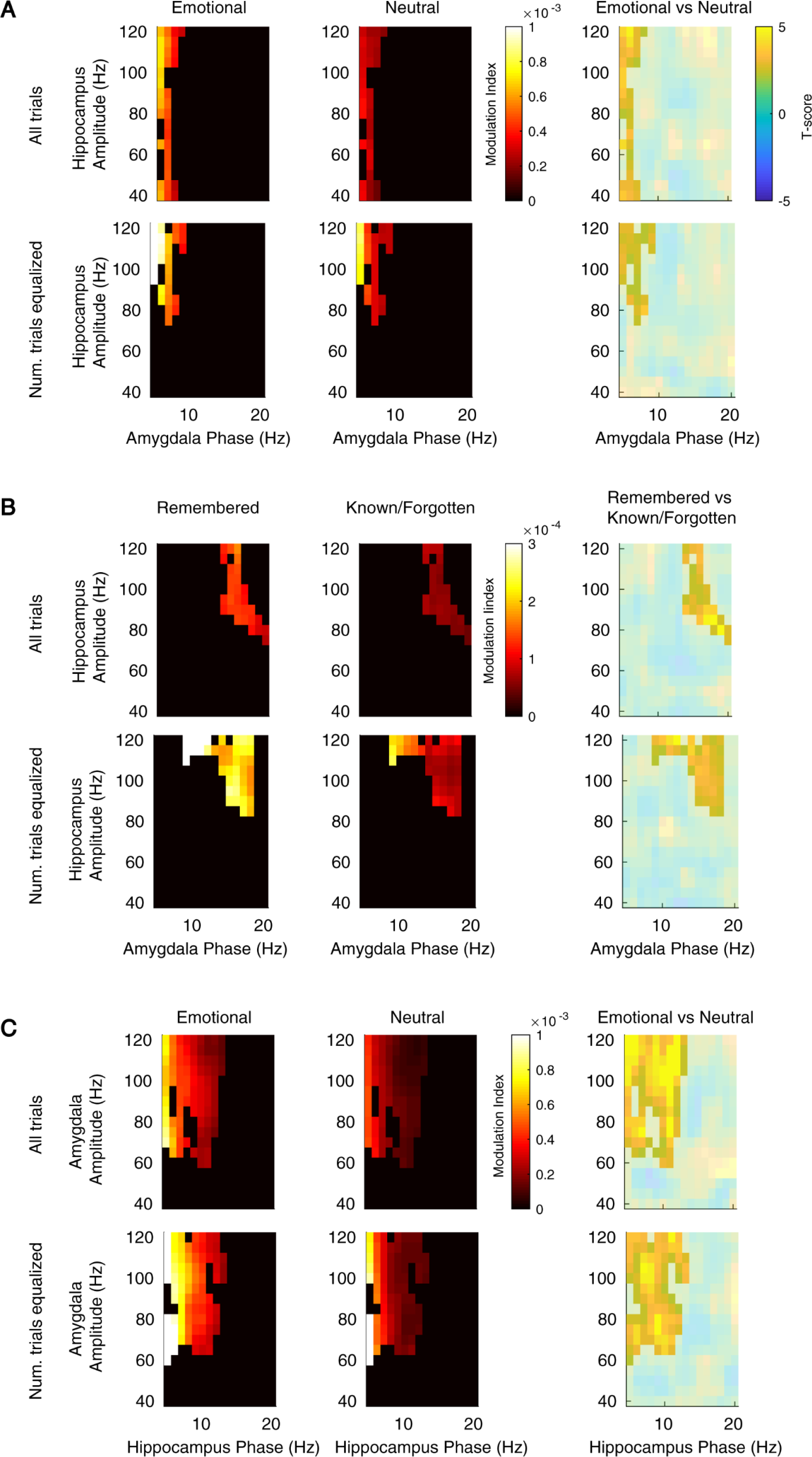
Between-region phase-to-amplitude coupling comodulograms (Cohort 1). (**A**) Amygdala phase (x-axes) to hippocampus amplitude (y-axes) coupling during emotional (top left) and neutral (top middle) conditions. Color code represents the modulation index (see Materials and Methods). Group-level statistics map (top right) ensuing from the main effect of emotion (eR+eKF vs. nR+nKF) contrast where color code represents the T-score (summed t value=104.95, *P*=9.99 x 10^-5^). All comodulograms were masked by a final alpha of 0.05 using two-sided nonparametric cluster-based permutation test. In this analysis all trials for each condition were included. Bottom row uses the same convention as top row but controlling the number of trials per condition (summed t value=73.70, *P*=0.0116) (**B**) Same conventions as in (**A**) but representing the main effect of memory (eR+nR vs. eKF+nKF) for amygdala phase to hippocampus amplitude (all trials, summed t values=89.87, *P*=0.017; number of trials equalized summed t value=127.51, *P*=0.0063). (**C**) Same figure conventions as (**A**) and (**B**) but showing the main effect of emotion for hippocampus phase to amygdala amplitude coupling (all trials, summed t value=347.55, *P*=9.99 x 10^-5^, number of trials equalized, summed t values=279.02, *P*=9.99 x 10^-5^, respectively). All reported main effects were computed including lateral bipolar channels from all patients with simultaneous hippocampus and amygdala recordings (*n=*8 patients, 9 amygdala-hippocampal electrode pairs) during 0.41–1.1 s post-stimulus onset.

**Fig. S13.**
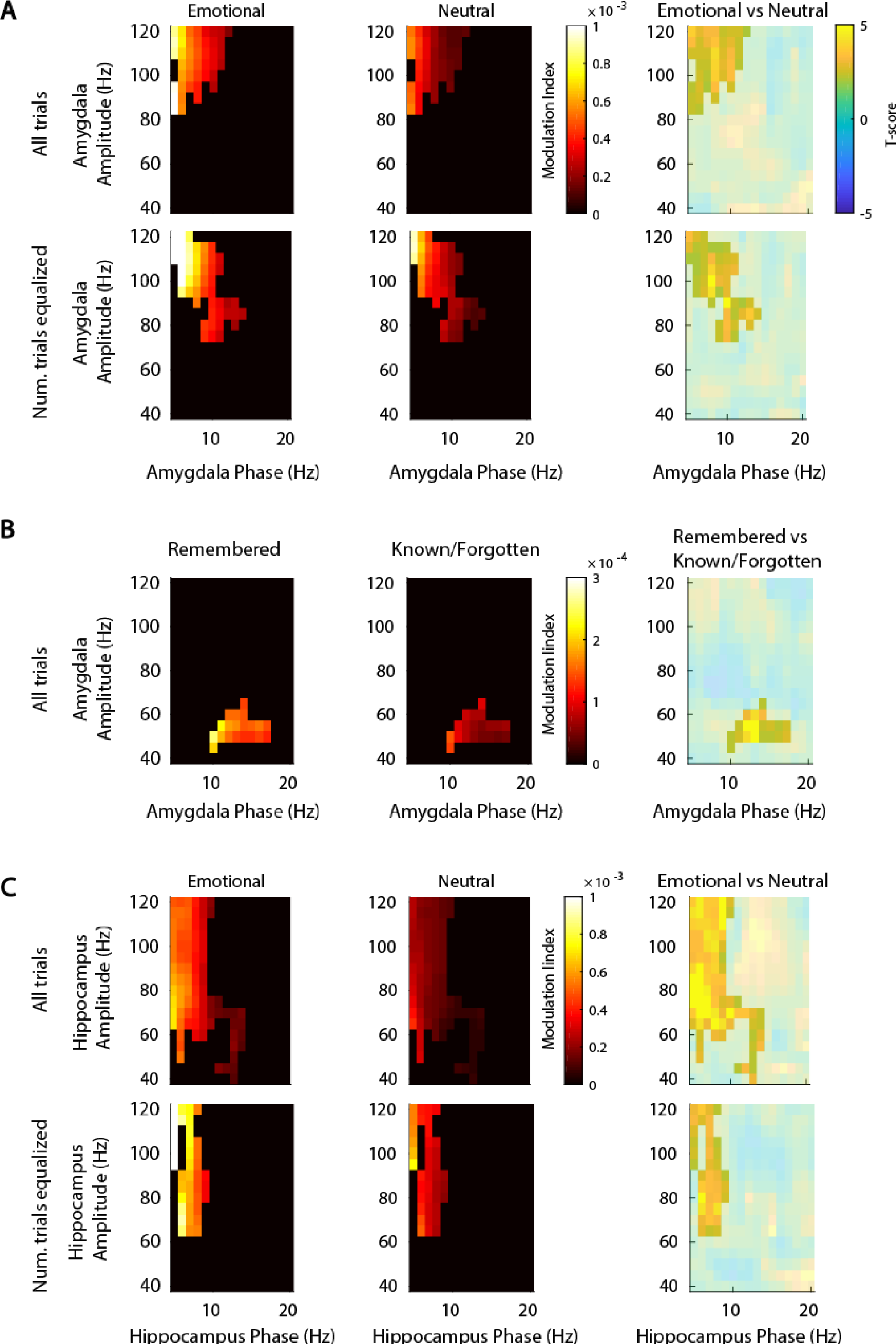
Within-region phase-to-amplitude coupling comodulograms (Cohort 1). (**A**) Amygdala phase (x-axes) to amygdala amplitude (y-axes) coupling during emotional (top left) and neutral (top middle) conditions. Color code represents the modulation index. Group-level statistics map (top right) ensuing from the main effect of emotion (eR+eKF vs. nR+nKF) contrast where color code represents the T-score (summed t value=121.44, *P*=0.0047). All comodulograms were masked by a final alpha of 0.05 using two-sided nonparametric cluster-based permutation test. In this analysis all trials for each condition were included. Bottom row uses the same convention as top row but controlling the number of trials per condition (summed t value=136.83, *P*=9.99 x 10^-5^). (**B**) Same conventions as in (**A**) but representing the main effect of memory (eR+nR *vs*. eKF+nKF) for amygdala phase to amygdala amplitude coupling (all trials, summed t value=63.13, *P*=0.024; not significant when number of trials were equalized). (**C**) Same figure conventions as in (**A**) but showing the main effect of emotion for hippocampus phase to hippocampus amplitude coupling (summed t value=329.19, *P*=9.99 x 10^-5^, summed t value=119.05, *P*=9.99 x 10^-5^, respectively). All reported main effects were computed including lateral bipolar channels from all patients with simultaneous hippocampus and amygdala recordings (*n=*8 patients, 9 amygdala-hippocampal electrode pairs) during 0.41–1.1 s post-stimulus onset.

**Fig. S14.**
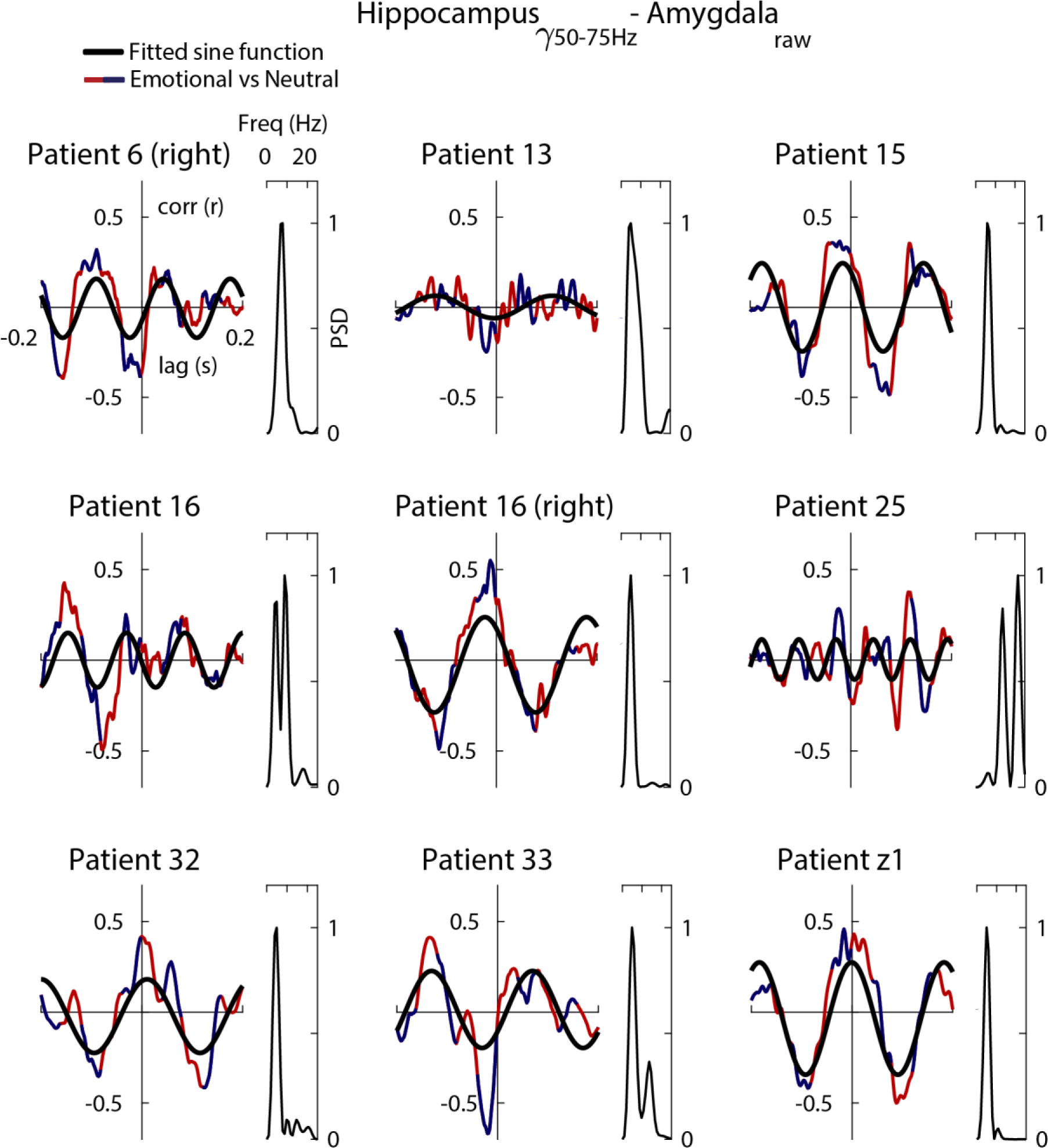
Inter-regional cross-correlation between emotional *vs*. neutral peak trigger average (PTA; blue/red) for each patient’s amygdala-hippocampus pair (Cohort 1). Each PTA was computed by first filtering hippocampus gamma activity (50–75 Hz), then identifying gamma peaks (minimum separation 0.1 s) and finally averaging the raw traces from amygdala recordings (± 0.12 s) centered (t=0 s) around the hippocampus gamma peaks (see Materials and Methods). Each subplot represents a lateral amygdala and lateral hippocampus bipolar channel pair. Right insert for each PTA: power spectral density (PSD) taken over the entire cross-correlogram to find the main spectral component that dominates the PTA. The PSD peak was used to fit the optimal sine wave (black), in the least squares sense. Note that all amygdala traces show a theta component reflected in the PSD and in the fitted (black) sine wave. All reported main effects were computed including lateral bipolar channels from all patients with simultaneous hippocampus and amygdala recordings (*n=*8 patients, 9 amygdala-hippocampal electrode pairs) during 0.41–1.1 s post-stimulus onset.

**Fig. S15.**
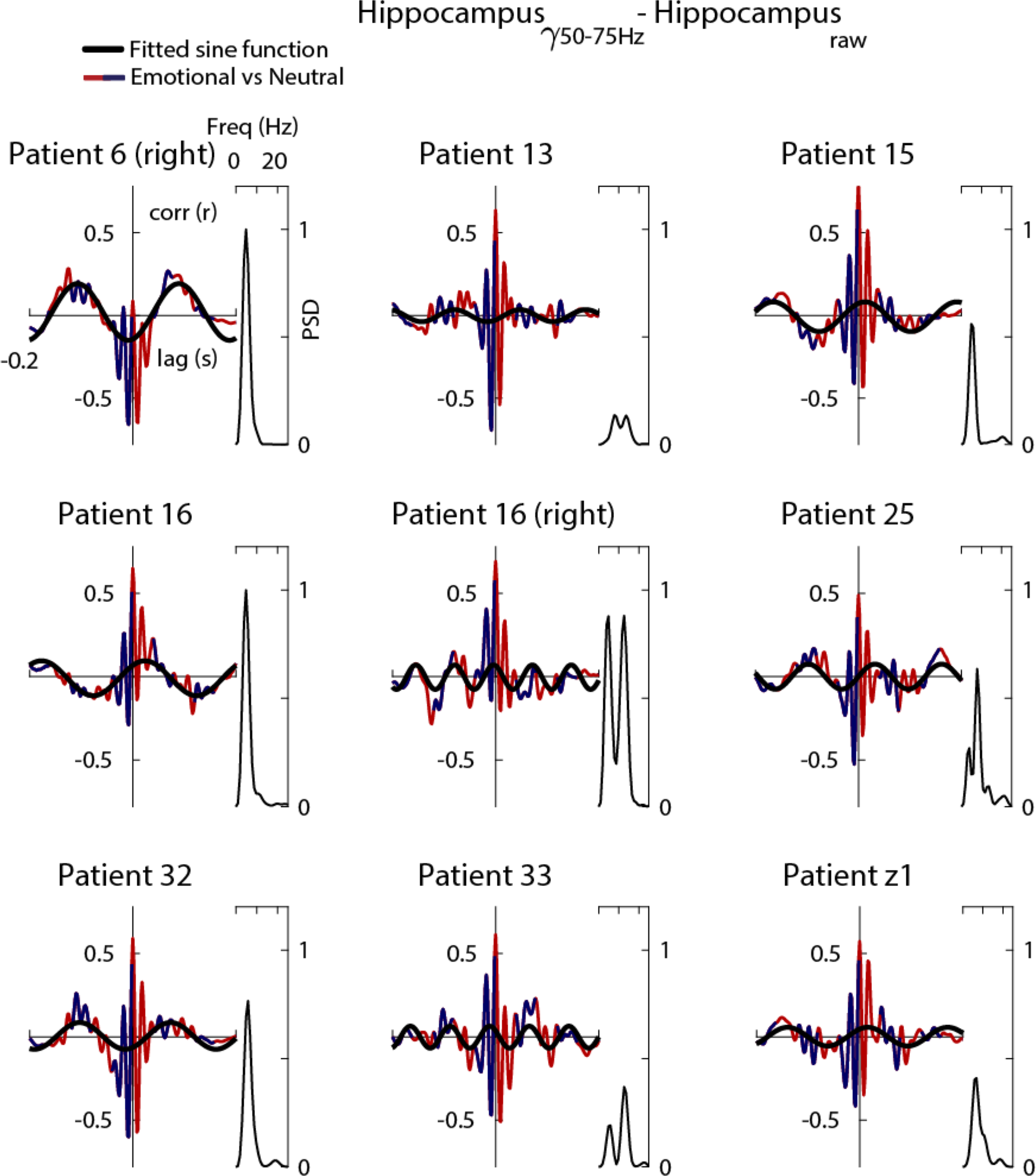
Single contact within-hippocampus cross-correlation between emotional vs. neutral peak trigger average (PTA; dark blue/dark red) (Cohort 1). Each PTA was computed by filtering hippocampus gamma activity (50–75 Hz), then identifying gamma peaks (minimum separation 0.1 s) and finally averaging the raw traces from the same hippocampus recordings (± 0.12 s) centered (t=0 s) around the peaks (see Materials and Methods). Each subplot represents a lateral hippocampus bipolar channel. Right insert: power spectral density (PSD) taken over the entire cross-correlogram to find the main spectral component that dominates the PTA. The PSD peak was used to fit the optimal sine wave (black), in the least squares sense. Note that all hippocampal traces show a theta component reflected in the PSD and in the fitted (black) sine wave. All reported main effects were computed including lateral hippocampal bipolar channels from all patients with simultaneous hippocampus and amygdala recordings (*n=*8 patients, 9 hippocampal electrodes) during 0.41–1.1 s post-stimulus onset.

**Fig. S16.**
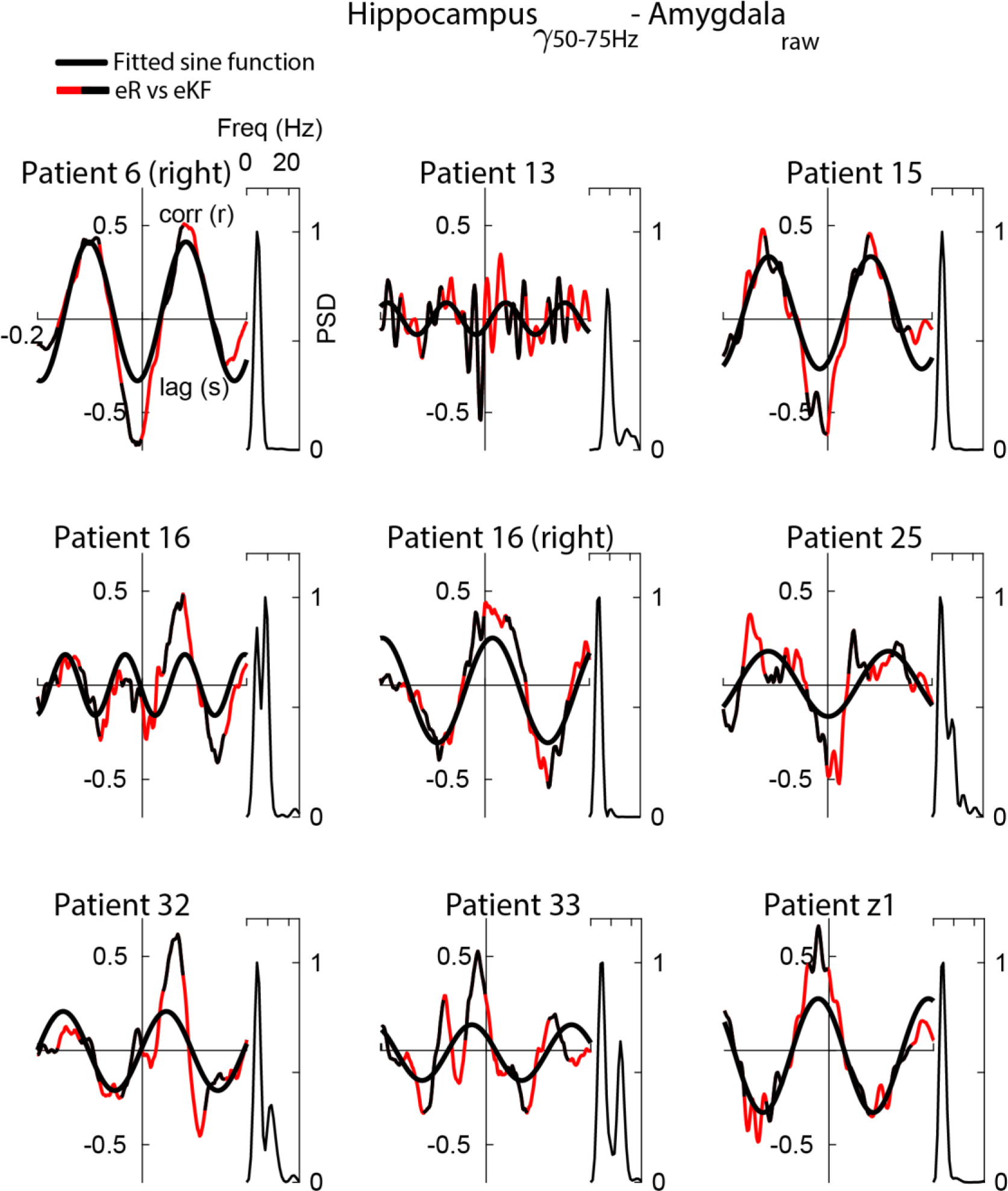
Inter-regional cross-correlation between emotionally remembered (eR) *vs*. emotionally known plus forgotten (eKF) peak trigger average (PTA; red/black) (Cohort 1). Each PTA was computed by filtering hippocampus gamma activity (50 –75 Hz), then identifying gamma peaks (minimum separation 0.1 s) and finally averagingthe raw traces from amygdala recordings (± 0.12 s) centered (t=0 s) around the hippocampus gamma peaks (see Materials and Methods). Each subplot represents a lateral amygdala and lateral hippocampus bipolar channel pair. Right insert: power spectral density (PSD) taken over the entire cross-correlogram to find the main spectral component that dominates the PTA. The PSD peak was used to fit the optimal sine wave (black), in the least squares sense. Note all amygdala traces show a theta component reflected in the PSD and in the fitted (black) sine wave. All reported simple effects (eR *vs*. eKF) were computed including lateral bipolar channels from all patients with simultaneous hippocampus and amygdala recordings (n=8 patients; 9 amygdala-hippocampal electrode pairs) during 0.41–1.1 s post-stimulus onset.

**Fig. S17.**
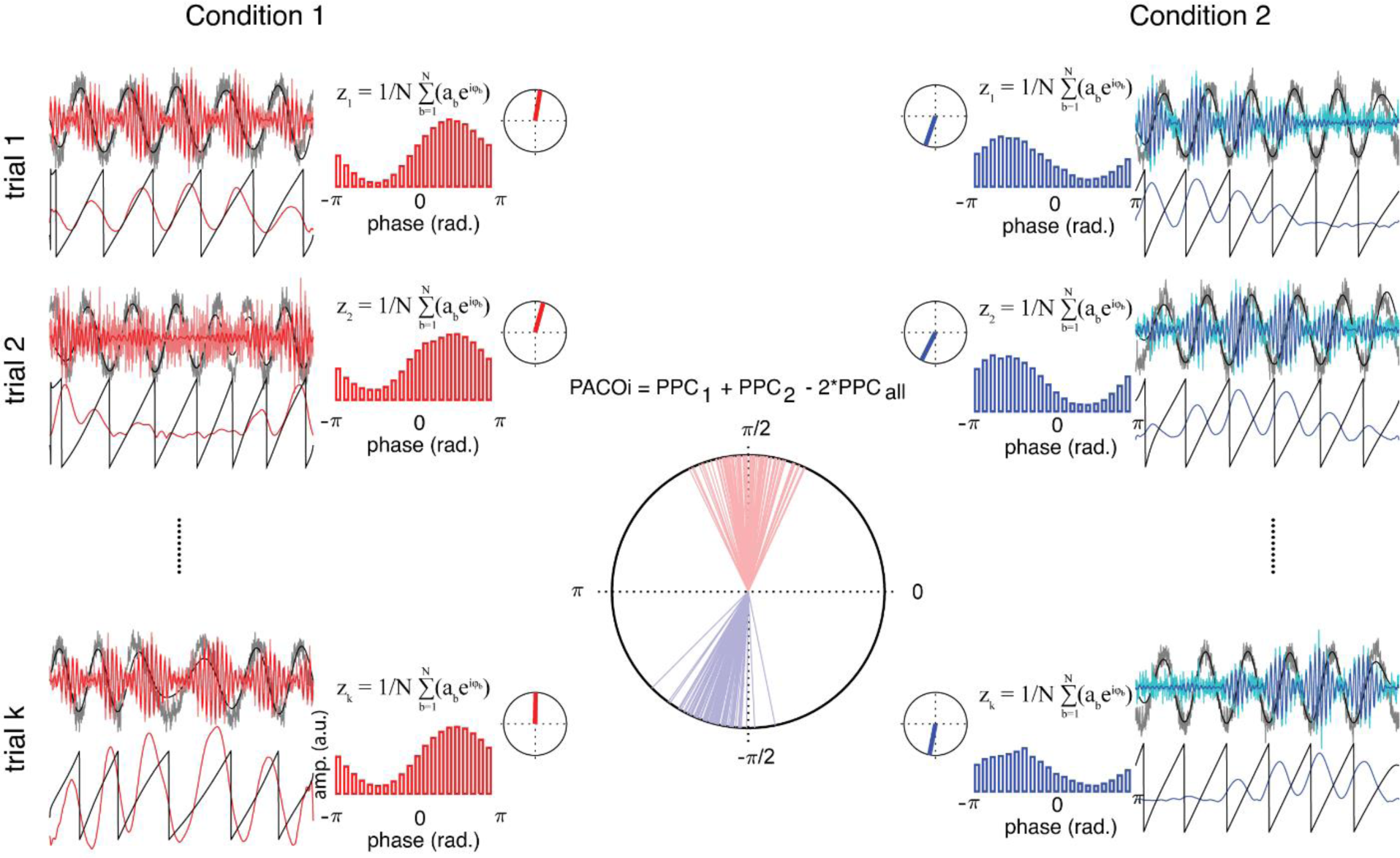
Phase-Amplitude Coupling Opposition index (PACOi). PACOi measures the phase opposition between two Phase-Amplitude Coupling (PAC) estimates obtained from two experimental conditions (left and right columns). The rationale of the metric is to test whether the high frequency amplitude of condition 1 *vs*. 2 locks at different phase bins. If PAC concentrates at different phase bins, the sum of the pairwise-phase consistency (PPC) of each experimental condition will exceed the PPC obtained from all trials together. Here we represent a simulation in which for each experimental condition and trial, the time series are filtered into low (left column; black traces) and high frequencies (left column: red traces) and the analytic phase and amplitude are computed taking the Hilbert transform respectively (for illustrative purposes we only show a few seconds of the time series). Second, the analytic phase of the low frequency is binned (b=20) and the high frequency amplitude is averaged within each phase bin, forming a histogram (left column; red histogram). Third, for each trial, this histogram is transformed into a complex number (zk; inserted formula) by multiplying the high frequency amplitude (histogram y-axis: ab) by the low frequency phase (histogram x-axis: exp(i^φb^); where φ_b_ denotes the average phase of each bin). This z value is a weighted average that indicates the phase of the lower-frequency oscillation at which the amplitude of the high-frequency oscillation is strongest. The z_k_ complex values were taken for each experimental trial and were normalized to unit length. The right column has the same conventions as the left but for condition 2. Middle column: The phase opposition is based on the sum of the PPC of each condition relative to the PPC calculated taking together the trials of the two conditions (PACOi=PPC_eR_+PPC_eKF_– 2*PPC_eR,eKF_). The PPC is defined as 

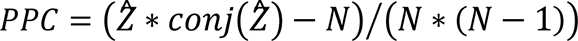

and 

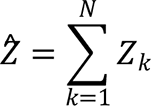

being the sum of single trial complex z values (N=total number of trials). PPC varies between 0 (random) and 1 (maximum pairwise phase consistency). Statistical significance was established by permuting (n=1000 times) the trials of each experimental conditions for a number of pair frequencies (5–20 Hz in 1 Hz steps and 40–120 Hz in 5 Hz steps). For each patient and bipolar channel, we selected the frequency phase-amplitude pairs that show a significant PACOi (uncorrected).

**Fig. S18.**
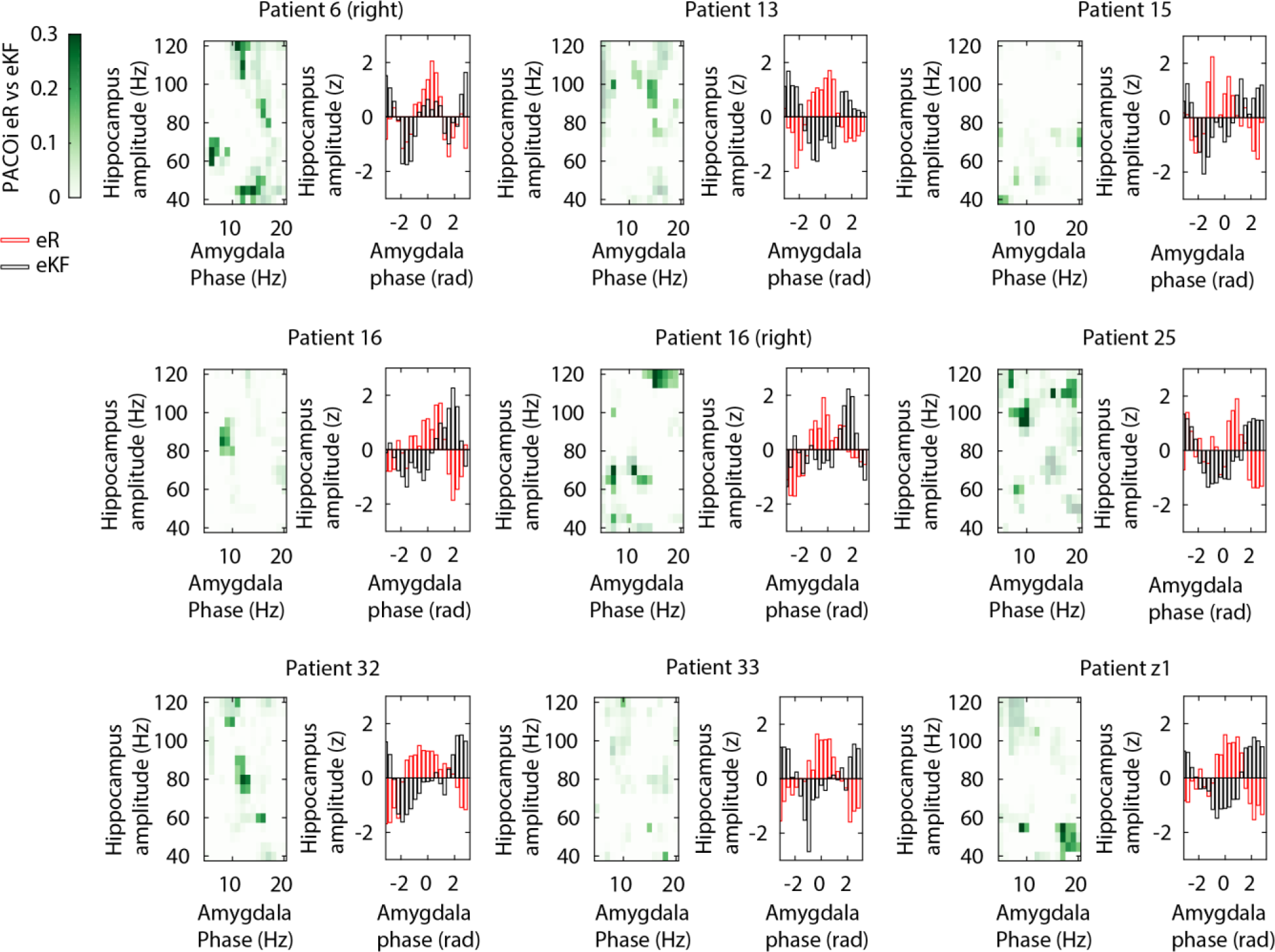
Phase-Amplitude Coupling Opposition index (PACOi) for aversive remembered and aversive known/forgotten trials for individual amygdala-hippocampal electrode pairs (Cohort 1). For each pair, left: comodulogram showing the amygdala phase (x-axes) to hippocampus amplitude (y-axes) phase opposition between emotional remembered (eR) *vs*. emotional know + forgotten (eKF) trials. Colorbar indicates the PACOi values (see Materials and Methods, fig. S13) masked by (uncorrected) statistical contrast between the eR *vs*. eKF group of trials. For each amygdala-hippocampal electrode pair, right: averaged hippocampus amplitude to amygdala phase histogram resulting from the significant (masked comodulogram) PACOi. Red represents eR and black eKF. PACOi analyses were performed over the 0.41–1.1 s post-stimulus time interval.

**Fig. S19.**
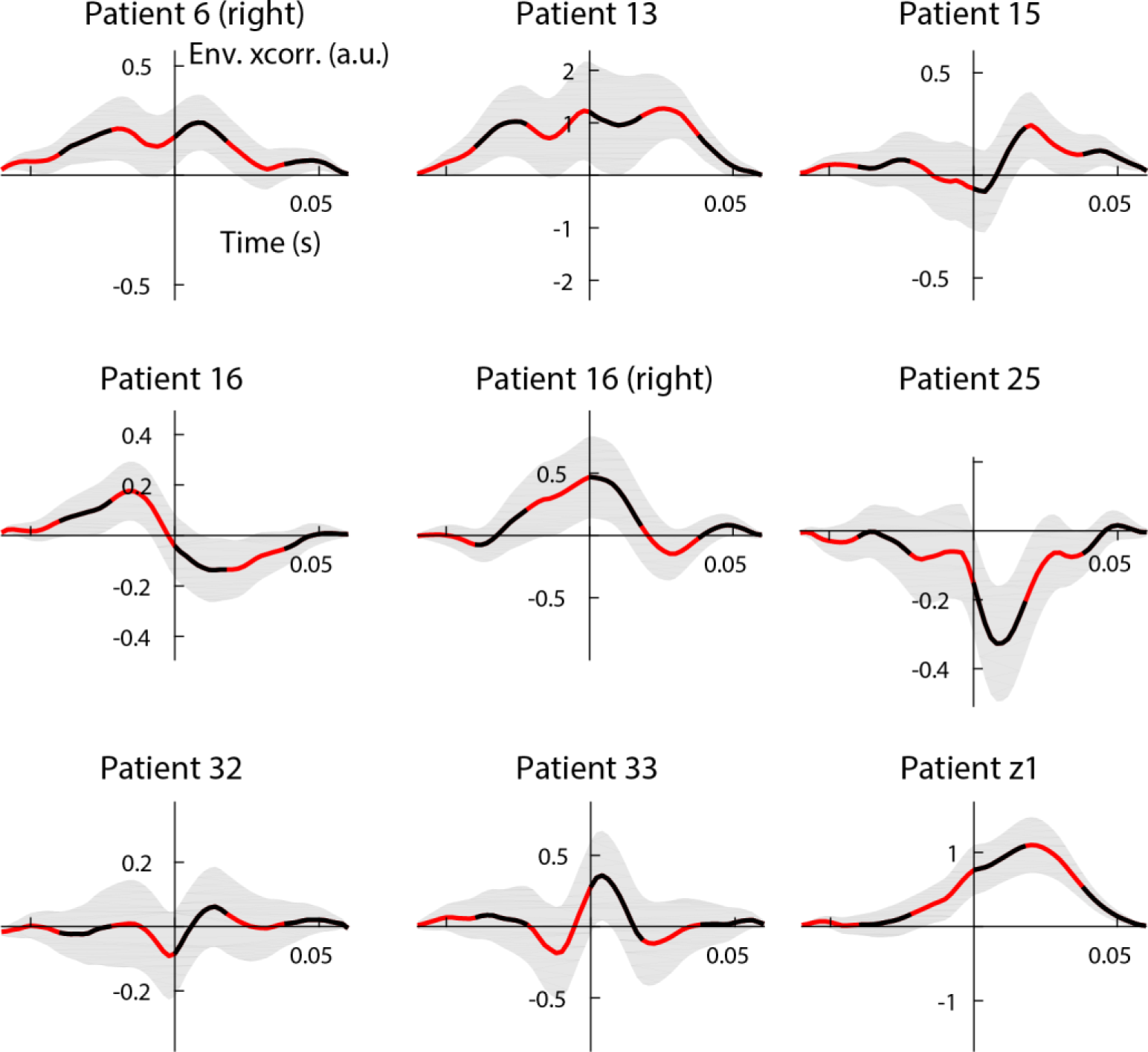
Broadband gamma (60–120 Hz) transient connectivity analysis between hippocampus and amygdala assessed by the amplitude envelope of cross-correlation (Cohort 1). Red/black curves represent the average amplitude envelope cross-correlation (Env. xcorr.) contrast between eR *vs*. eKF (see fig. S18). The input signals to compute the cross-correlation were the hippocampus and amygdala broadband gamma activities taken over the 0.41–1.1 s post-stimulus time interval. Shaded grey area represents the s.e.m. taken over the epochs, a.u.: arbitrary units.

**Fig S20.**
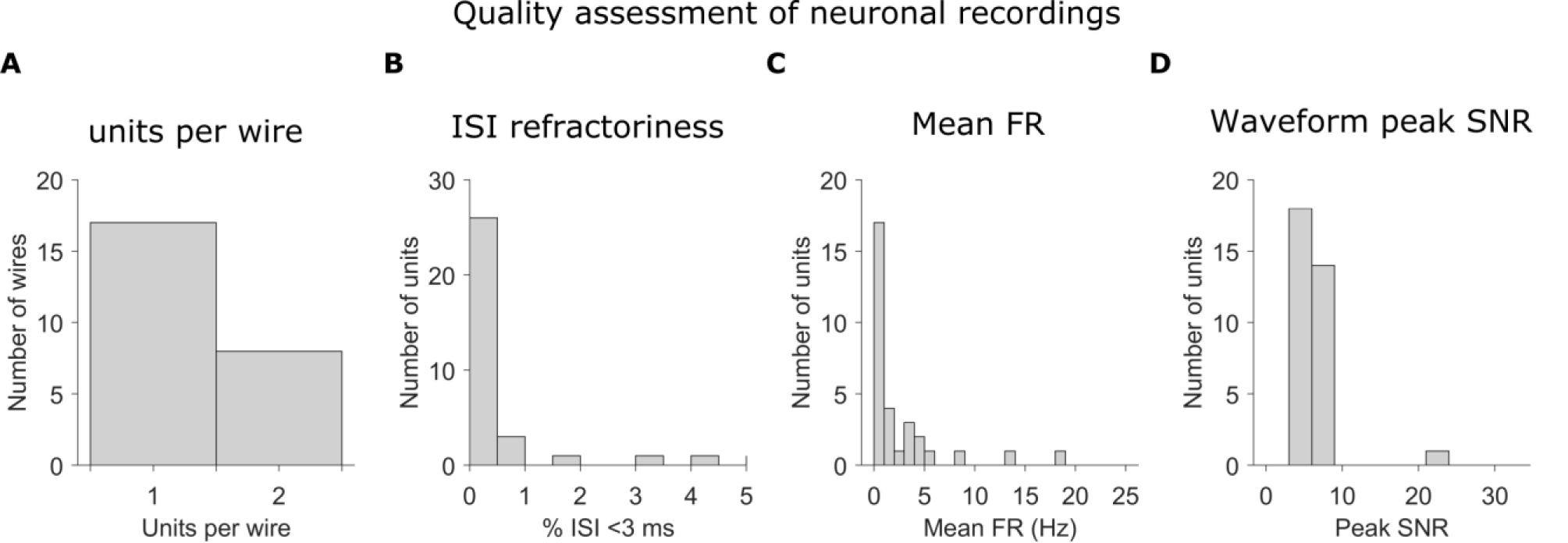
Quality assessment of neuronal recordings and spike sorting (Cohort 2). We recorded *n*=33 neurons from the hippocampus. **(A)** Histogram of how many neurons were identified on each wire. On average, 1.32 ± 0.09 neurons per wire (mean ± s.e.m) were recorded. Only wires with at least one neuron are counted. **(B)** Histogram of the proportion of inter-spike interval (ISIs) shorter than 3 ms. On average, units exhibited 0.65 ± 0.27% ISIs that were shorter than 3 ms (mean ± s.e.m). The majority of units had less than 1% of short ISIs. **(C)** Histogram of the mean firing rates over the 33 neurons recorded in the hippocampus. On average, neurons exhibited mean FRs of 4.53 ± 1.54 spikes/s (mean ± s.e.m). The firing rates of these neurons did not differ significantly between aversive scenes that were later remembered (FRs eR = 4.32 ± 1.40 spikes/s, mean ± s.e.m) or forgotten (FRs eKF = 4.36 ± 1.45 spikes/s, mean ± s.e.m). **(D)** Waveform peak signal-to-noise ratio (SNR) for each neuron. On average, the SNR of the mean waveform peak was 6.50 ± 0.49 (mean ± s.e.m).

**Fig S21.**
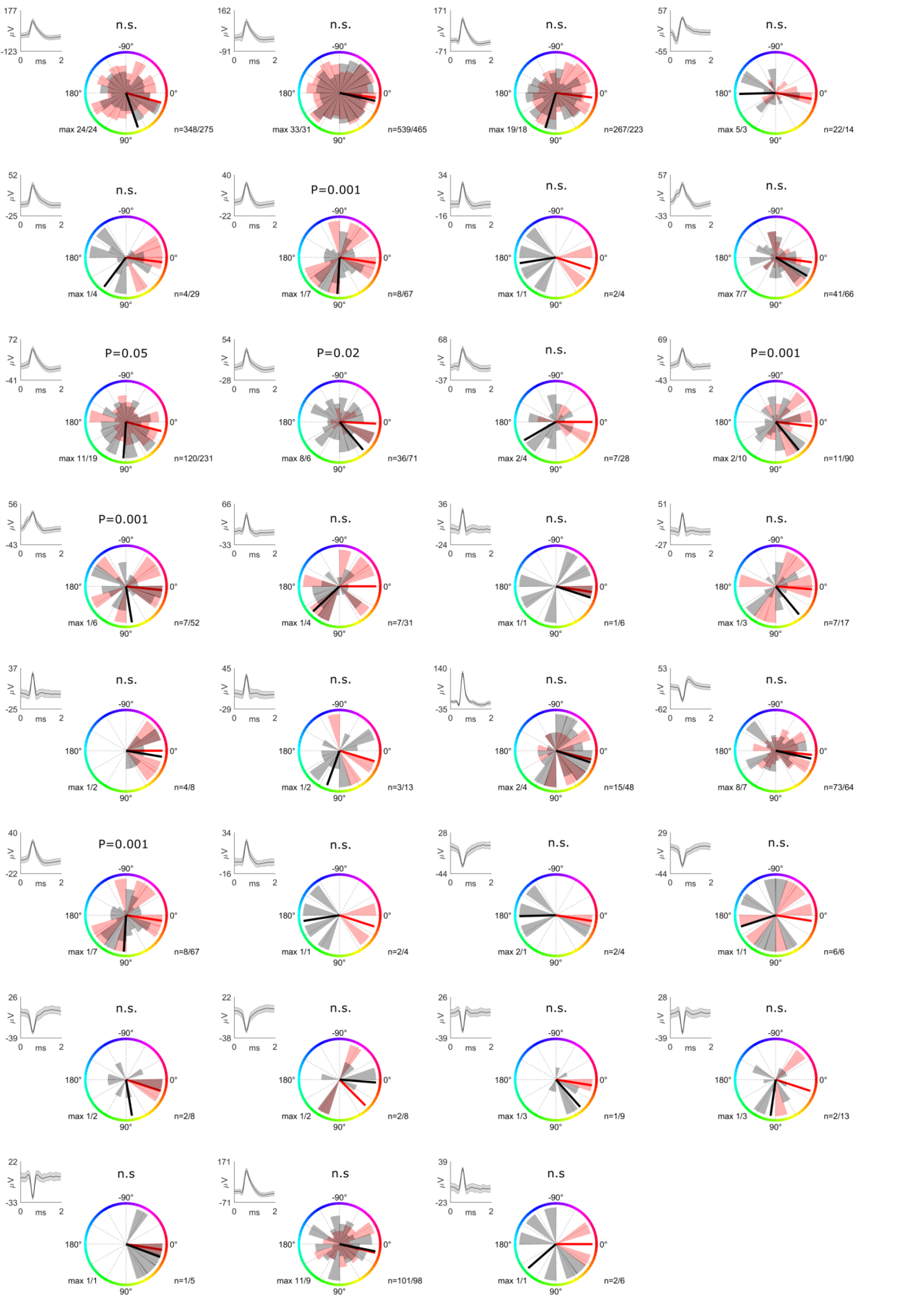
Single neuron SFC amygdala theta hippocampus spikes *n*=31 (Cohort 2). SFC phase opposition between eR and eKF trials for each observed neuron. Red and black shaded area represents spikes per phase bin (n=20) for each condition, red and black line is the realigned preferential phase for eR and eKF condition, respectively. *P* values from the circular Kuiper test are reported on the top of each neuron showing significant phase opposition (n.s.: not significant); max is the maximum number of spikes occurring in a certain bin for each condition (eR/eKF); *n* is the number of total spikes per neuron observed from 0.4 to 1.1 s for each condition (eR/eKF). Upper left subpanel: single-neuron waveform (mean ± std).

**Table S1.** Cohort 1 patient demographic and clinical data

| Patient | Sex | Hand-<br>edness | Age (year<br>range) | Aetiology | Lesion location | VIQ | PIQ | % trials without epileptic<br>spikes in amygdala |  | % trials without<br>epileptic spikes in<br>hippocampus |  |
| --- | --- | --- | --- | --- | --- | --- | --- | --- | --- | --- | --- |
| **Z1** | F | R | 50-57 | Hippocampal sclerosis | Left hippocampal<br>sclerosis | 75 | 98 | 87.71 % |  | 87.71 % |  |
| **02** | F | R | 34-41 | Hippocampal sclerosis<br>plus focal dysplasia | Right hippocampal<br>sclerosis plus<br>porencephalic cyst over<br>the parieto-occipital<br>junction | 103 | 91 | 92.43 % |  | NA |  |
| **04** | F | R | 18-25 | Focal dysplasia | Extensive lesion over<br>the left frontal region<br>involving dorsolateral<br>and orbitofrontal cortex<br>and anterior border of<br>the cingulum | 86 | 80 | 87.5 % |  | NA |  |
| **06** | M | R | 42-49 | Hippocampal sclerosis | Left hippocampal<br>sclerosis | 115 | 86 | Left 98.3 % | Right 100 % | Right 100 % |  |
| **13** | F | R | 42-49 | Focal dysplasia | Right basal temporal<br>cortex | 87 | 88 | 77.3% |  | 77.3% |  |
| **15** | F | R | 34-41 | Focal dysplasia | Left temporal pole | 97 | 98 | 100 % |  | 100 % |  |
| **16** | M | R | 26-33 | Reactive gliosis, diffuse<br>microglia activation and<br>small vessel<br>vasculopathy | Medial wall of the left<br>parietal region<br>(precuneus and<br>posterior cingulum) | 102 | 93 | Left 94.06% | Right 88.98% | Left 94.06% | Right 88.98% |
| **21** | F | R | 26-33 | Periventricular<br>heterotopia | Left occipital horn<br>heterotopia | 104 | 87 | Left 90% | Right 94.11% | NA |  |
| **25** | M | R | 26-33 | Focal dysplasia | Right posterior<br>temporobasal region | 116 | 97 | 79.16% |  | 79.16% |  |
| **27** | M | R | 18-25 | Focal dysplasia | Left temporal pole | 69 | 102 | 98.33% |  | NA |  |
| **32** | M | R | 58-65 | Encephalocele | Left temporal pole | 100 | 119 | Left 100 % | Right 100 % | Left 100% |  |
| **33** | F | R | 18-25 | Inflammatory lesion | Right parietal cortex | 78 | 93 | 96.58% |  | NA |  |
| **34** | M | R | 18-25 | Posttraumatic | Left temporo-occipital | 86 | 81 | 100% |  | NA |  |

Abbreviations: PIQ/VIQ=Procedural/Verbal Intelligence Quotient of the Wechsler Adult Intelligence Scale, version III (WAIS-III);

**Table S2.** Cohort 2 patient demographic, clinical data

| Patient | Sex | Hand-<br>edness | Age (year<br>range) | Aetiology | Lesion<br>location |
| --- | --- | --- | --- | --- | --- |
| Z1 | F | R | 50-57 | Hippocampal<br>sclerosis | Left<br>hippocampus |
| Z2 | M | R | 34-41 | Focal cortical<br>dysplasia | Right frontal<br>cortex |
| Z5 | M | R | 26-33 | Dysembryoplastic<br>neuroepithelia<br>tumor | Left anterior<br>hippocampus |
| Z6 | F | R | 26-33 | Unclear etiology | Unclear |
| Z8 | F | R | 26-33 | Non-lesional |  |
| Z10 | M | R | 50-57 | Hippocampal<br>sclerosis | Right<br>hippocampus |

**Table S3.**
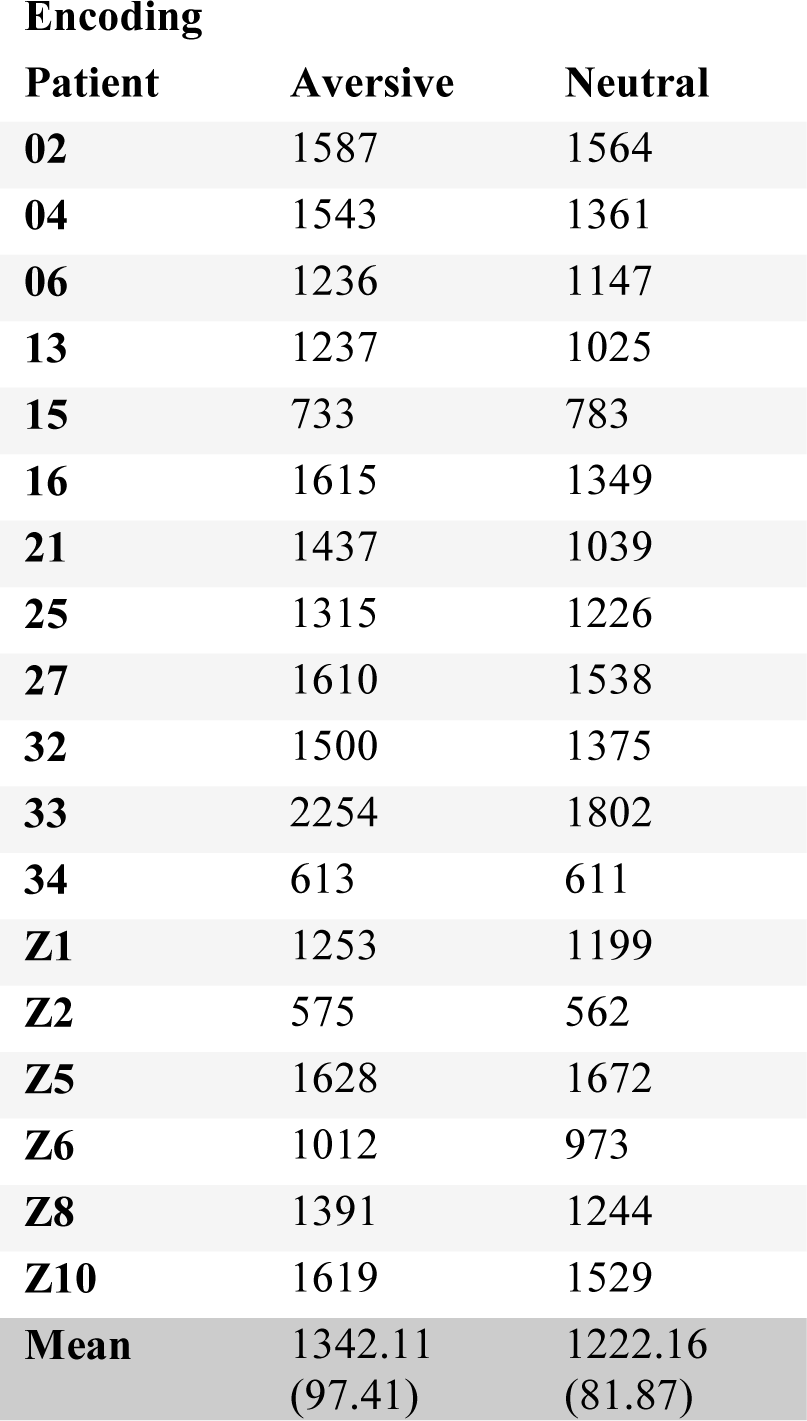
Behavioral data from 18 patients (Cohorts 1 and 2). Reaction time at encoding. Single patient RT values, group mean ± s.e.m in ms. RTs are longer in response to aversive *vs*. neutral pictures (paired t-test T_17_ =3.66, P=0.002).

**Table S4.** Behavioral data from 18 patients (Cohorts 1 and 2). Reaction time at recognition for all memory categories: remember (R), remember false alarm (RFA), know (K), know false alarm (KFA), miss (Miss) and correct rejected (Crj) for emotional (e) and neutral (n) trials. Single patient RT values, group mean and s.e.m in ms. We compared RTs of remembered (R) items to the RTs of know (K) and correct rejection (Crj) responses for emotional (eR, eK, Crje) and neutral items (nR, nK, Crjn). The ANOVA revealed a significant main effect of memory F_(2,16)_=8.85, *P*=0.003, but no main effect of emotion F_(1,17)_=0.48, *P*=0.49 or emotion by memory interaction F_(2,16)_=2.87, *P*=0.086, η^2^=0.26.

| **Recognition** |  |  |  |  |  |  |
| --- | --- | --- | --- | --- | --- | --- |
| **Patient** | **Re** | **R FAe** | **Ke** | **K FAe** | **Misse** | **Crje** |
| **02** | 1562 | 1858 | 1787 | 1859 | 1776 | 1737 |
| **04** | 1647 | 1566 | 2348 | 1833 | 2057 | 1855 |
| **06** | 1381 | 2160 | 1697 | 2000 | 1426 | 1432 |
| **13** | 1368 | 1259 | 1735 | 2219 | 1772 | 1611 |
| **15** | 1524 | 1462 | 1702 | 1554 | 1868 | 1859 |
| **16** | 1587 | 1845 | 2125 | 2454 | 2208 | 1700 |
| **21** | 1453 | 1269 | 1845 | 2502 | 2066 | 1850 |
| **25** | 1957 | 1899 | 2203 | 2565 | 1781 | 2129 |
| **27** | 1546 | 1392 | 2146 | 2377 | 2390 | 2398 |
| **32** | 1483 | 1597 | 1770 | 1788 | 1880 | 1690 |
| **33** | 1482 | 1487 | 1927 | 1653 | 1902 | 1421 |
| **34** | 698 | 701 | 975 | 907 | 925 | 839 |
| **Z1** | 1283 | 1475 | 2108 | 2317 | 1831 | 1949 |
| **Z2** | 554 | 598 | -- | -- | 549 | 506 |
| **Z5** | 1684 | 1543 | 2095 | 2316 | 1976 | 2042 |
| **Z6** | 1378 | 1687 | 1391 | 1461 | 1186 | 1293 |
| **Z8** | 1518 | 1834 | 1726 | 1717 | 1695 | 1781 |
| **Z10** | 1567 | 1577 | 3188 | 2245 | 1566 | 1343 |
| **Mean** | 1426.22<br>(77.11) | 1511.61<br>(91.91) | 1927.52<br>(108.92) | 1986.29<br>(105.44) | 1714.11<br>(106.74) | 1635.27<br>(106.52) |

|  | **Rn** | **R FAn** | **Kn** | **K FAn** | **Missn** | **Crjn** |
| --- | --- | --- | --- | --- | --- | --- |
| **02** | 1592 | -- | 1749 | 1797 | 1539 | 1454 |
| **04** | 1527 | 1887 | 1947 | 2113 | 1622 | 1742 |
| **06** | 1436 | 1986 | 1783 | 1758 | 1183 | 1185 |
| **13** | 1792 | 1509 | 1885 | 2026 | 1450 | 1464 |
| **15** | 1643 | 1501 | 1473 | 1518 | 1579 | 1499 |
| **16** | 1665 | 2303 | 2557 | 2690 | 1836 | 1881 |
| **21** | 1186 | 1095 | 1769 | 1897 | 1510 | 1617 |
| **25** | 2104 | 2009 | 2275 | 2397 | 1827 | 1884 |
| **27** | 1686 | 2035 | 2136 | 2273 | 2415 | 1975 |
| **32** | 1420 | 1721 | 1814 | 1882 | 1649 | 1651 |
| **33** | 1573 | 1465 | 1755 | 1393 | 1159 | 1055 |
| **34** | 759 | 715 | 894 | 805 | 760 | 800 |
| **Z1** | 3116 | 1442 | 1803 | 1921 | 1831 | 1949 |
| **Z2** | 583 | 590 | 834 | -- | 546 | 539 |
| **Z5** | 1881 | 1708 | 2131 | 2339 | 2062 | 1920 |
| **Z6** | 1185 | 1223 | 1571 | 1495 | 1293 | 1252 |
| **Z8** | 1632 | 2468 | 1766 | 1645 | 1638 | 1485 |
| **Z10** | 1134 | 2096 | 1663 | 3095 | 1356 | 1197 |
| **Mean** | 1550.77<br>(127.91) | 16325.29<br>(122.92) | 1766.94<br>(98.51) | 1943.76<br>(125.30) | 1499.22<br>(101.96) | 1449.88<br>(92.39) |

**Table S5.** Total number of trials per patient and condition (Cohort 1). r, right electrode in the case of bilateral electrodes.

| **Patient** | **eR**<br>**aversive**<br>**remember** | **eK**<br>**aversive**<br>**know** | **eF**<br>**aversive**<br>**forgotten** | **nR**<br>**neutral**<br>**remember** | **nK**<br>**neutral**<br>**know** | **nF**<br>**neutral**<br>**forgotten** |
| --- | --- | --- | --- | --- | --- | --- |
| **06r** | 10 | 13 | 17 | 11 | 25 | 43 |
| **13** | 19 | 7 | 14 | 19 | 16 | 44 |
| **15** | 19 | 8 | 13 | 23 | 21 | 36 |
| **16** | 24 | 6 | 10 | 48 | 12 | 18 |
| **16r** | 24 | 6 | 10 | 48 | 12 | 18 |
| **25** | 17 | 14 | 9 | 32 | 9 | 39 |
| **32** | 12 | 14 | 14 | 39 | 18 | 23 |
| **33** | 21 | 12 | 5 | 20 | 12 | 47 |
| **Z1r** | 21 | 11 | 6 | 4 | 21 | 51 |

**Table S6.** Number of analyzed trials per patient and conditions following exclusions based on both time- and frequency-domain artifact rejection, and random selection used for the connectivity analysis (Cohort 1). NA, not applicable (data from patient Z1 were not used in testing for emotion by memory interaction due to low number of nR trials).

| **Patient** | **eR**<br>**aversive**<br>**remember** | **eKF**<br>**aversive**<br>**know/forgotten** | **nR**<br>**neutral**<br>**remember** | **nKF**<br>**neutral**<br>**know/forgotten** | **Random**<br>**selection**<br>**of trials** |
| --- | --- | --- | --- | --- | --- |
| **06r** | 10 | 29 | 11 | 68 | 10 |
| **13** | 14 | 14 | 14 | 35 | 14 |
| **15** | 19 | 19 | 22 | 57 | 19 |
| **16** | 21 | 16 | 43 | 26 | 16 |
| **16r** | 19 | 11 | 33 | 22 | 11 |
| **25** | 14 | 9 | 15 | 26 | 9 |
| **32** | 12 | 28 | 39 | 41 | 12 |
| **33** | 14 | 10 | 16 | 37 | 10 |
| **Z1r** | 16 | 11 | 2 | 52 | 2/NA |

## Acknowledgments

We thank the electroencephalography technicians at the Hospital Ruber Internacional, as well as Isabel Montón Quesada and Linda Zhang for technical assistance.

## Author contributions

B.A.S. designed the experiment. M.C., C.M.-B., M.Y., J.S. and R.T. collected data. M.C., D.L.-S., S.M., C.O, L.K., N.A. and B.A.S. performed analyses. R.T. and A.G.-N. monitored patients and performed clinical evaluation. L.S. performed surgical electrode implantation. M.C., D.L.-S., and B.A.S. wrote the paper with input from all of the other authors.

## Data and materials availability

All data needed to evaluate the conclusions in the paper are present in the paper and/or the supplementary materials. Analysis code will be made publicly available after publication.

